# Parental stress and family quality of life in families of individuals living with Angelman syndrome

**DOI:** 10.1101/2025.05.07.25327165

**Authors:** Catherine Merton, Angela Gwaltney, Anna Booman, Sarah Nelson Potter, Anne C. Wheeler, Rene L. Barbieri-Welge, Lucia T. Horowitz, Rachel J. Hundley, Lynne M. Bird, Wen-Hann Tan, Anjali Sadhwani

**Affiliations:** Division of Genetics and Genomics, Boston Children’s Hospital, Boston, MA; RTI International, Research Triangle Park, NC; Developmental Evaluation Clinic, Rady Children’s Hospital San Diego, San Diego, CA; Greenwood Genetic Center, Greenwood, SC; Division of Developmental Medicine, Vanderbilt University School of Medicine, Nashville, TN; University of California, San Diego, Department of Pediatrics and Rady Children’s Hospital San Diego Genetics / Dysmorphology, San Diego, CA; Department of Psychiatry and Behavioral Sciences, Boston Children’s Hospital, Boston, MA; Harvard Medical School, Boston, MA

**Keywords:** Parental stress, psychosocial stress, family interaction, developmental disabilities

## Abstract

**Background:** Angelman syndrome (AS) is a developmental disorder caused by one of four molecular etiologies. Affected individuals have intellectual disability (ID), limited speech, seizures, and sleep problems. Parents of individuals with AS exhibit elevated stress compared to parents of individuals with other IDs. We examined parental stress and family quality of life (FQOL) over time in families of individuals living with AS.

**Methods:** Data were collected in a natural history study of AS. The Parenting Stress Index, Third Edition (PSI) and the Beach Center FQOL scale assessed parent stress and FQOL. Stress and FQOL were examined across AS molecular subtypes, and predictors were analyzed using a generalised linear model. Relationships between parental stress and FQOL were examined using Pearson correlations and a stepwise mixed-linear model approach.

**Results:** Our sample consisted of 231 families of individuals living with AS. Parental stress was clinically elevated and was highest in families of individuals with *UBE3A* mutations, while FQOL did not differ across subtype in most domains. Increasing age predicted a decrease in parental stress but did not predict FQOL. Elevated parental stress was additionally predicted by maladaptive behaviours and child male sex, while lower FQOL was predicted by child male sex, parent marital status, and family income. Parental stress had a small negative impact on FQOL.

**Conclusions:** Stress is elevated in parents of individuals with AS across subtypes and has a small negative impact on family quality of life. Interventions to reduce stress have potential to improve individual and family well-being.

## BACKGROUND

Angelman syndrome (AS) is a rare neurogenetic disorder caused by loss of expression of *UBE3A*, which may be due to a deletion of the AS critical region on maternally-inherited chromosome 15q11-q13 (65%-70%) (deletion subtype); paternal uniparental disomy (UPD) for chromosome 15q11-q13 (5%-10% of cases), imprinting center defects (ImpD) (5%-10%), or *UBE3A* pathogenic variants (10%-15%) (non-deletion subtypes).^1^ Deletions are further subclassified based on their size as class I (5.9 Mb, 40%), class II (5.0 Mb, 53%), or atypical (7%).^1^ AS is characterised by severe intellectual disability (ID), lack of speech, seizures, and sleep difficulties.^2^ At six years of age, AS individuals exhibit cognitive developmental age between 15 and 27 months; individuals with a deletion subtype have more severe impairments than individuals with a non-deletion subtype.^3^ Maladaptive behaviours, including hyperactivity, irritability, and aggression, are common, especially among individuals with non-deletion subtypes.^4,5^ Sleep difficulties (increased sleep latency and frequent nocturnal awakenings) and epilepsy are reported in ∼90% of individuals with AS.^1,6^ Communication, sleep, and behaviour challenges negatively impact the quality of life for the individuals and their caregivers, and increase family burden.^7^ Treatment is currently limited to symptomatic interventions, although promising disease-modifying therapies are in development.^8^

Parents of individuals with AS report elevated stress compared to parents of neurotypical individuals and individuals with other IDs and autism.^9–12^ A previous study comparing parental stress across AS molecular subtypes found that clinically elevated stress was more common among non-deletion subtypes, although parents of individuals with *UBE3A* pathogenic variants were not included.^13^ Other studies support an association between behaviour and sleep difficulties and parental stress, but the impact of impaired cognitive and adaptive functioning, epilepsy, and other medical complications is unknown.^5,12–14^ In the general population, demographic and psychosocial factors, including child sex, maternal education, family income, race, and ethnicity may impact parental stress.^15,16^ Health-related child quality of life (QoL) is shown to be lower in individuals with AS than neurotypical individuals,^17–19^ but less is known about family quality of life (FQOL) in families of individuals living with AS. Further, little is known about how parental stress changes over time or across different age groups, and the relationship between parental stress and FQOL has not yet been described.

This study sought to examine predictors of, and associations between, parental stress and FQOL across age and molecular subtypes in AS. The aims of this study were:

1. Describe parental stress and FQOL in families of individuals living with AS across molecular subtype and over time.
2. Describe predictors of parental stress and FQOL.
3. Analyze how parental stress correlates with FQOL.

## METHODS

### Participants

Participants were enrolled in the AS Natural History Study (NHS) (ClinicalTrials.gov identifier: NCT00296764) at one of six study sites: Rady Children’s Hospital-San Diego; Texas Children’s Hospital; Greenwood Genetics Center; Boston Children’s Hospital; Vanderbilt University Medical Center; and Cincinnati Children’s Hospital Medical Center between 2006 and 2014. Participants underwent annual evaluations of cognition, adaptive functioning, and behaviour. Included participants had genetics testing results specifying a molecular subtype of AS (deletion class 1, deletion class 2, UPD, ImpD, or *UBE3A* pathogenic variant). Data collected for participants 22 years of age and over were excluded from this analysis. Age 22 was selected since it is the age at which individuals with disabilities lose eligibility for special education services in the state of the lead study site.^20^ In instances where parent-reported measures were completed by different caregivers, we included measures completed by the caregiver who attended the majority of visits. The term “parent” will be used to refer to a participant’s primary caregiver.

### Ethics approval

The NHS was approved by institutional review boards of each participating institution. Informed consent was obtained from a legally authorised representative for each participant.

### Measures

#### Use of standardised measures outside age range

Some measures to assess parental stress and development are standardised for use in children of specific ages. However, these measures were administered to individuals of all ages in this study because the developmental age of all the participants in this study fell within the normative age range for which these measures were developed.^3^

#### Parental Stress and FQOL

Parental stress was assessed using the Parenting Stress Index, Third Edition (PSI).^21^ The PSI is a parent-completed measure with 120 items in three domains: child characteristics, parent characteristics, and situational/demographic life stress. The Child Domain includes six subdomains: Distractibility/Hyperactivity, Adaptability, Reinforces Parent, Demandingness, Mood, and Acceptability. The Parent Domain includes seven subdomains: Competence, Isolation, Attachment, Health, Role Restriction, Depression, and Spouse/Parenting Partner Relationship. The Life Stress Domain does not contribute to the Total Stress score and was excluded from this study. Parents rate items on a five-point scale (strongly agree, agree, not sure, disagree, strongly disagree), and domain scores and a total score are generated. Higher scores indicate higher parental stress; scores above the 85^th^ percentile indicate clinically significant stress. The PSI has been reported in several AS studies.^12–14^ It is standardised for use among parents of children aged one month to 12 years.

FQOL was assessed using the Beach Center Family Quality of Life Scale (FQOL Scale).^22^ The FQOL Scale is a parent-report measure that assesses the importance of, and satisfaction with, five domains of quality of life: Family Interaction, Parenting, Emotional Well-Being, Physical/Material Well-Being, and Disability-Related Support. Only the satisfaction scale was utilised and the importance scale was excluded, consistent with prior studies.^23–25^ Parents rate 25 items on a five-point scale (very dissatisfied, satisfied, neither, satisfied, very satisfied), with higher scores indicating higher levels of satisfaction. Scores are calculated as the mean of each subscale.^26^ The FQOL Scale has been used in studies of individuals with developmental disorders,^23,27,28^ and in one other AS study.^5^

#### Development, Adaptive, and Behavioural Functioning

Development and adaptive functioning were assessed with the Bayley Scales of Infant and Toddler Development, Third Edition (BSID-III)^29^ and the Vineland Adaptive Behaviour Scales, Second Edition (VABS-II),^30^ respectively. The BSID-III assesses cognitive, receptive and expressive language, and fine and gross motor skills using standardised activities administered by a trained rater. Raw and age equivalent (AE) scores are generated for each domain. This measure is standardised for use in children up to 42 months. The VABS-II is a clinician-administered parent interview that assesses adaptive functioning in three domains: communication, daily living skills, and socialization; for individuals under the age of seven years, a fourth domain (motor skills) is also assessed. The motor skills domain was included for all individuals in this study. Domain and AE scores, as well as an overall Adaptive Behaviour Composite (VABS-II composite) score, are generated.

Maladaptive behaviours were assessed using the Aberrant Behaviour Checklist-Community version (ABC-C).^31^ This measure has 58-items rated on a scale of zero to three, with zero indicating a behaviour is not a problem to three indicating a behaviour is a severe problem. The ABC-C has five domains: Irritability, Agitation, Crying; Lethargy and Social Withdrawal; Stereotypic Behaviour; Hyperactivity and Noncompliance; and Inappropriate Speech.^31^ The Inappropriate Speech domain was excluded from this study since the majority of participants in the study are nonverbal. Previous studies have shown significant correlation between ABC-C Irritability and Hyperactivity scores and parental stress levels, while ABC-C Stereotypy and Lethargy scores have weak associations with parental stress.^5^ Therefore, only the Irritability and Hyperactivity domains were analyzed. The ABC-C has been used in several previous studies of individuals with AS.^4,5,32^

#### Demographic and Medical Characteristics

Demographic data were collected at baseline. A structured medical history was collected at each visit to assess seizures and other medical problems. Participants’ sleep habits were assessed via a parent interview by study investigators.

### Statistical methods

#### Participant Data and Description of PSI and FQOL scores

Descriptive analyses were performed on demographic variables, baseline medical and developmental variables, and baseline PSI and FQOL Scale scores. Baseline visits are defined as the first visit at which the PSI and/or FQOL Scales were completed. The percentage of respondents with PSI Total Stress and Child and Parent Domain stress scores above the 85^th^ percentile was calculated. Characteristics across subtypes were compared using Pearson Chi-square, Fisher’s exact, or one-way analysis of variance (ANOVA) tests; if significant, post hoc Tukey’s Honestly Significant Difference (HSD) test was conducted to determine which subtype groups were significantly different.

#### Predictors of PSI and FQOL scores

We investigated predictors of PSI Child Domain stress, Parent Domain stress, Total Stress, and FQOL subscales using generalised linear models (GLM), with adjustments for repeated measures in order to account for within-subject correlation. This was achieved by modeling the subject effect as part of the error structure rather than as a fixed effect. For ease of interpretation, age was centered at six years. Molecular subtype, participant sex, age at visit, race, ethnicity, annual household income, number of persons in household, number of siblings, maternal education, parent marital status, seizure severity, average number of hospitalizations per year, frequency of nocturnal awakenings, VABS-II Composite, and BSID-III Cognitive AE were analyzed as predictors. Variables were individually entered into a bivariate model with either PSI or FQOL score as the outcome. Only the significant terms in the bivariate models were then included in the multivariate model to determine if they were still predictors of PSI/FQOL scores when controlling for all other significant terms in the bivariate model. For models with only one significant term, the multivariate model was the same as the bivariate model. To ensure comparability between the bivariate and multivariate models, the bivariate analyses were restricted to data from the subset of observations with complete data on all covariates included in the multivariate model. Post hoc analyses were performed on significant categorical variables to identify differences in least squares means with Bonferroni adjustments for multiple comparisons by subtype. To analyze predictors of FQOL, a log link function was used to account for a non-normal distribution of FQOL scores and predicted coefficients were exponentiated to rescale to the original scale.

#### Relationship between PSI and FQOL scores

Multiple analyses were conducted to investigate how parental stress contributes to FQOL. PSI Parent Domain, Child Domain, and Total Stress scores were included in the GLM when analyzing predictors of FQOL, with adjustments for repeated measures, described above.

Pearson correlations were calculated to analyze the relationship between baseline PSI Domain scores and FQOL subscale scores. Finally, a stepwise, generalised linear-mixed model approach was used to investigate the effects of PSI Child and Parent Domains on FQOL subscales with adjustments for repeated measures. Similar to previous analyses, age was centered at six years, a log link function was used, and predicted coefficients were exponentiated to rescale to the original scale. Starting with the unconditional model (a model with no predictors), various covariates, interaction terms, and random effects were added and subtracted using the lowest Bayesian information criterion (BIC) to determine the best model for predicting each FQOL subscale. Covariates were molecular subtype, age, ethnicity, parent marital status, seizure severity, VABS-II Composite, BSID-III Cognitive AE, ABC-C Irritability and Hyperactivity scores, molecular subtype by age interaction, and molecular subtype by seizure severity interaction. Although age was not statistically significant, age was included in the conditional models as a covariate to explicitly test the effect of time in addition to the effect of molecular subtype.

SAS (version 7.15; SAS Institute, Cary, NC) was used for all analyses; *p*<.05 was considered statistically significant.

## RESULTS

### Demographics

Data from 231 participants who completed the PSI during study participation were analyzed (Table 1). Of those, 214 completed the FQOL scale. Participants had an average of two to three annual evaluations. The majority of participants were non-Hispanic White, and parents were married or in marriage-like relationships. No significant differences in demographic parameters were observed across subtypes except for parent marital status: parents of individuals with ImpD reported highest rates of marriage/marriage-like relationships while parents of individuals with Class I deletions reported lowest rates of marriage/marriage-like relationships (*p*<.001). Medical and developmental covariates are presented in Table 2. Significant differences across subtype were observed for seizure severity (*p*<.001), VABS-II composite score (*p* = .004), ABC-C Irritability score (*p*<.001), and BSID-III Cognitive age equivalent score (*p*<.001), consistent with previous research.^3,5,33,34^

**Table 1.** Demographic Characteristics at Baseline Visit (*n*=231)

| | Class I deletion<br>($n = 62$)<br>26.8% | Class II deletion<br>($n = 92$)<br>39.8% | *UBE3A*<br>pathogenic<br>variant<br>($n = 28$)<br>12.1% | ImpD<br>($n = 21$)<br>9.1% | UPD<br>($n = 28$)<br>12.1% | p-value |
| --- | --- | --- | --- | --- | --- | --- |
| **Sex, $n$ (%)** |  |  |  |  |  | .95 |
| **Female** | 34 (54.8%) | 47 (51.1%) | 14 (50.0%) | 10 (47.6%) | 16 (57.1%) |  |
| **Male** | 28 (45.2%) | 45 (48.9%) | 14 (50.0%) | 11 (52.4%) | 12 (42.9%) |  |
| **Child age at baseline visit** |  |  |  |  |  |  |
| **Mean ($SD$)** | 4.6 (3.8) | 4.9 (3.9) | 5.4 (4.1) | 5.4 (4.2) | 6.8 (4.5) | .16 |
| **Range** | 1-17 | 1-20 | 1-15 | 2-21 | 2-21 |  |
| **Child age at final visit** |  |  |  |  |  |  |
| **Mean ($SD$)** | 7.1 (4.8) | 7.2 (4.4) | 7.4 (4.3) | 7.8 (4.2) | 8.3 (4.5) | .75 |
| **Range** | 1-21 | 1-21 | 1-19 | 3-21 | 2-21 |  |
| **Race ($n$ (%) White)** | 54 (87.1%) | 82 (89.1%) | 28 (100.0%) | 20 (95.2%) | 26 (92.9%) | .30 |
| **Ethnicity ($n$ (%) Hispanic/Latinx)** | 13 (21.0%) | 16 (17.4%) | 0 (0%) | 2 (9.5%) | 5 (17.9%) | .06 |
| **Annual Household Income, $n$ (%)** |  |  |  |  |  | .39 |
| **Under \$25,000** | 6 (9.7%) | 7 (7.6%) | 1 (3.6%) | 1 (4.8%) | 2 (7.1%) | |
| **\$25,000-\$49,999** | 13 (21.0%) | 14 (15.2%) | 4 (14.3%) | 1 (4.8%) | 1 (3.6%) | |
| **\$50,000-\$99,999** | 19 (30.7%) | 29 (31.5%) | 10 (35.7%) | 5 (23.8%) | 10 (35.7%) | |
| **\$100,000 or more** | 18 (29.0%) | 33 (35.9%) | 8 (28.6%) | 12 (57.1%) | 13 (46.4%) | |
| **Declined/Missing** | 6 (9.7%) | 9 (9.8%) | 5 (17.9%) | 2 (9.5%) | 2 (7.1%) |  |
| **# of persons supported on income, $n$ (%)** |  |  |  |  |  | .12 |
| **2** | 2 (3.2%) | 3 (3.3%) | 1 (3.6%) | 0 (0%) | 1 (3.6%) |  |
| **3** | 23 (37.1%) | 17 (18.5%) | 6 (21.4%) | 6 (28.6%) | 4 (14.3%) |  |
| **4** | 22 (35.5%) | 35 (38.0%) | 10 (35.7%) | 9 (42.9%) | 10 (35.7%) |  |
| **5** | 10 (16.1%) | 20 (21.7%) | 5 (17.9%) | 4 (19.1%) | 7 (25.0%) |  |
| **6 or more** | 4 (6.5%) | 10 (10.9%) | 1 (3.6%) | 1 (4.8%) | 4 (14.3%) |  |
| **Declined/Missing** | 1 (1.6%) | 7 (7.6%) | 5 (17.8%) | 1 (4.6%) | 2 (7.1%) |  |
| **# of persons in household, Mean ($SD$)** | 4.0 (1.2) | 4.4 (1.2) | 3.9 (0.9) | 4.1 (1.0) | 4.2 (1.0) | .60 |
| **# of siblings, Mean ($SD$)** | 1.4 (1.2) | 1.7 (1.2) | 1.1 (0.9) | 1.8 (1.9) | 1.8 (0.1) | 1.00 |
| **Respondent Relation, $n$ (%)** |  |  |  |  |  | .35 |
| **Mother** | 47 (75.8%) | 74 (80.4%) | 23 (82.1%) | 16 (76.2%) | 22 (78.6%) |  |
| **Father** | 7 (11.3%) | 7 (7.6%) | 1 (3.6%) | 4 (19.1%) | 5 (17.9%) |  |
| **Other** | 1 (1.6%) | 0 (0%) | 0 (0%) | 0 (0%) | 0 (0%) |  |
| **Maternal Education, *n* (%)** |  |  |  |  |  | .53 |
| **Less than college** | 27 (43.6%) | 38 (41.8%) | 6 (28.6%) | 6 (28.6%) | 12 (46.2%) |  |
| **College or more** | 35 (56.5%) | 53 (58.2%) | 15 (71.4%) | 15 (71.4%) | 14 (53.9%) |  |
| **Declined/Missing** | 0 (0%) | 1 (1.1%) | 7 (25.0%) | 0 (0%) | 2 (7.1%) |  |
| **Parent Marital Status, *n* (%)** |  |  |  |  |  | <.001 |
| **Single, never married** | 5 (8.1%) | 5 (5.4%) | 0 (0%) | 0 (0%) | 0 (0%) |  |
| **Married/Living in marriage-like relationship** | 45 (75.6%) | 74 (80.4%) | 23 (82.1%) | 19 (90.5%) | 24 (85.7%) |  |
| **Separated/Divorced** | 6 (9.7%) | 8 (8.7%) | 1 (3.6%) | . | 2 (7.1%) |  |
| **Declined/Missing** | 6 (9.7%) | 5 (5.4%) | 4 (14.6%) | 2 (9.5%) | 2 (7.1%) |  |

ImpD: Imprinting Center Defect; UPD: Uniparental Disomy

**Table 2.**
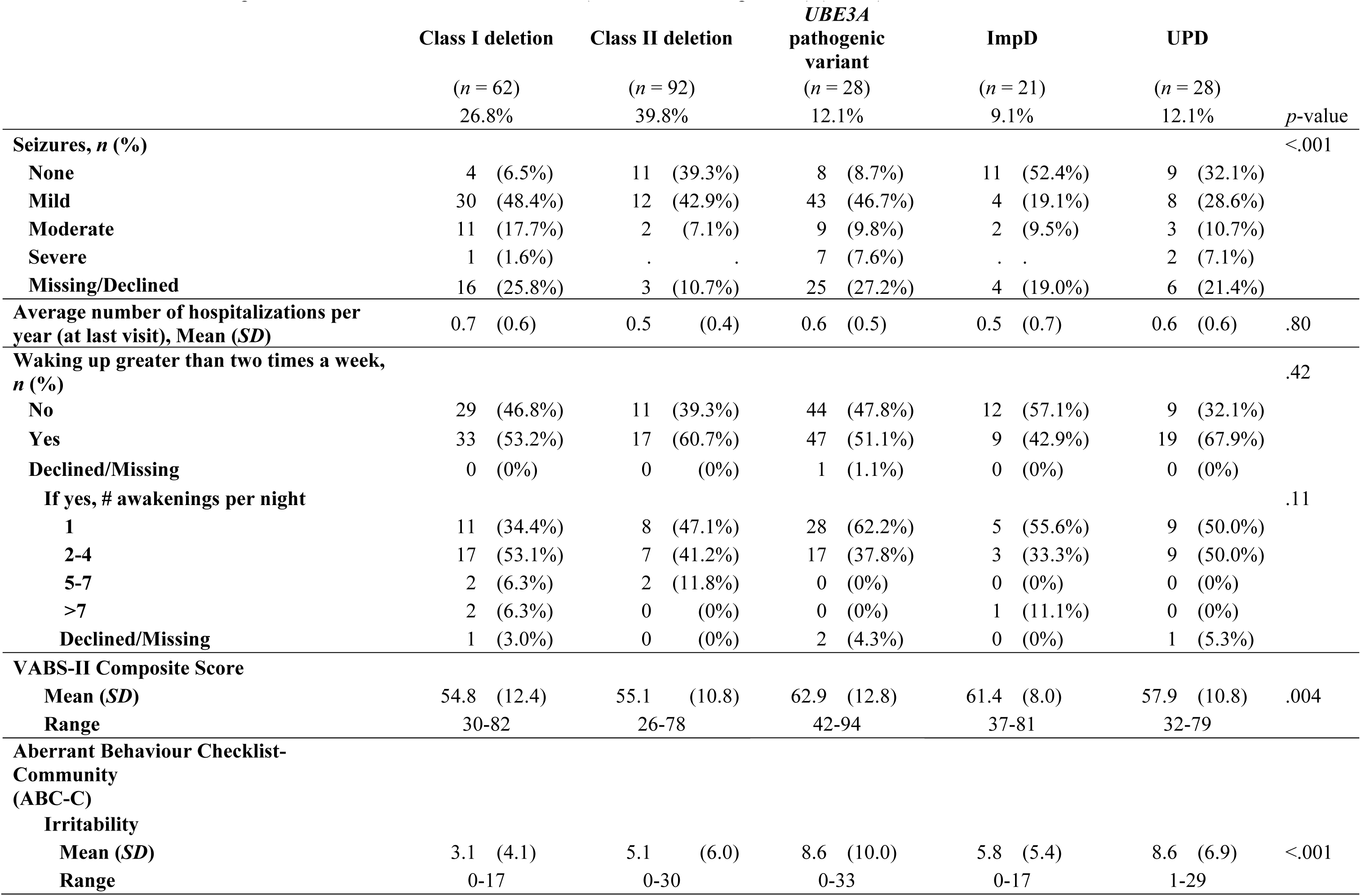
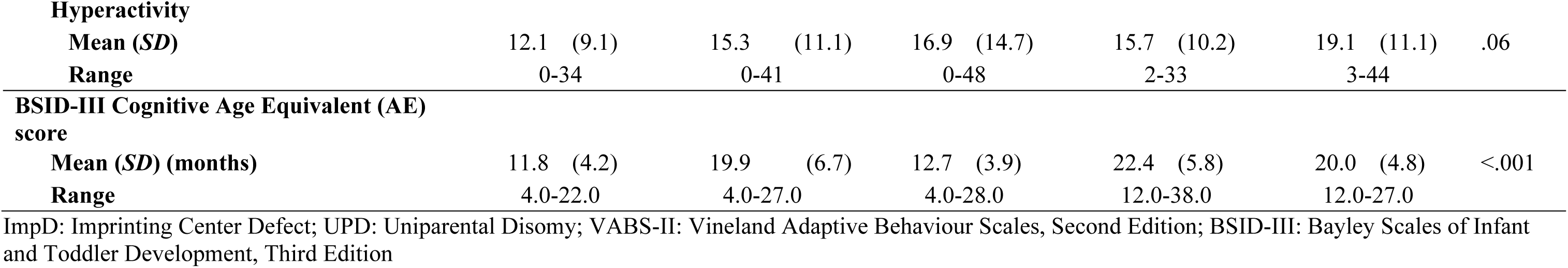
Medical and Developmental Characteristics at Baseline Visit (unless otherwise specified) (*n*=231)

### Description of PSI and FQOL across subtypes and over time

Table 3 summarises baseline PSI scores across subtypes. The percentage of respondents with Total Stress scores above the 85^th^ percentile, representing clinical significance, ranged from 19% in the class II deletion group to 48% in the *UBE3A* pathogenic variant group. Forty to 60 percent of respondents reported clinically significant Child Domain stress and 11 to 26 percent of respondents reported clinically significant Parent Domain stress. Table 4 summarises baseline FQOL scores across molecular subtype. Total mean score ranged from 3.7 to 4.1.

**Table 3.** Parenting Stress Index (PSI) Scores at Baseline Visit (*n*=231)

|  | Class I deletion |  |  | Class II deletion |  |  | UBE3A pathogenic variant |  |  | ImpD |  |  | UPD |  |  | *p*-value |
| --- | --- | --- | --- | --- | --- | --- | --- | --- | --- | --- | --- | --- | --- | --- | --- | --- |
|  | Mean | (SD) | % | Mean | (SD) | % | Mean | (SD) | % | Mean | (SD) | % | Mean | (SD) | % |  |
| **Total Stress (101-505)** | 231.3 | (35.1) | 21 | 226.3 | (34.4) | 19 | 262.0 | (47.5) | 48 | 243.1 | (32.3) | 30 | 242.6 | (33.5) | 28 | <.001 |
| **Child Domain** |  |  |  |  |  |  |  |  |  |  |  |  |  |  |  |  |
| **Total Child Domain Score (47-235)** | 115.0 | (18.3) | 45 | 114.0 | (17.9) | 40 | 128.5 | (25.6) | 59 | 123.1 | (17.6) | 60 | 123.0 | (18.6) | 56 | .003 |
| Distractibility/Hyperactivity (9-45) | 31.6 | (4.8) |  | 31.9 | (5.4) |  | 32.2 | (6.8) |  | 32.8 | (6.4) |  | 32.8 | (5.0) |  | .86 |
| Adaptability (11-55) | 26.0 | (7.0) |  | 26.3 | (5.9) |  | 29.3 | (6.8) |  | 30.0 | (5.2) |  | 30.0 | (7.8) |  | .005 |
| Reinforces Parent (6-30) | 8.7 | (3.0) |  | 8.5 | (2.8) |  | 9.6 | (3.8) |  | 9.1 | (2.8) |  | 7.8 | (1.8) |  | .24 |
| Demandingness (9-45) | 22.7 | (5.9) |  | 22.2 | (5.9) |  | 27.8 | (7.9) |  | 24.9 | (6.2) |  | 26.2 | (6.3) |  | <.001 |
| Mood (5-25) | 7.7 | (2.7) |  | 7.8 | (2.5) |  | 10.5 | (3.7) |  | 8.3 | (2.5) |  | 8.5 | (3.1) |  | <.001 |
| Acceptability (7-35) | 18.4 | (3.0) |  | 17.3 | (3.5) |  | 19.1 | (3.9) |  | 18.1 | (4.1) |  | 18.8 | (4.0) |  | .11 |
| **Parent Domain** |  |  |  |  |  |  |  |  |  |  |  |  |  |  |  |  |
| **Total Parent Domain Score (54-270)** | 116.4 | (22.5) | 12 | 112.7 | (22.3) | 11 | 133.4 | (25.6) | 26 | 120 | (22.1) | 20 | 120.3 | (22.2) | 15 | 0.002 |
| Competence (13-65) | 24.9 | (5.5) |  | 24.9 | (6.4) |  | 28.4 | (6.0) |  | 24.8 | (4.9) |  | 27.2 | (5.3) |  | .03 |
| Isolation (6-30) | 12.6 | (4.3) |  | 12.6 | (3.5) |  | 14.5 | (4.1) |  | 13.6 | (4.5) |  | 13.8 | (4.3) |  | .15 |
| Attachment (7-35) | 11.2 | (3.2) |  | 11.3 | (3.0) |  | 12.1 | (3.3) |  | 11.3 | (2.6) |  | 11 | (2.0) |  | .69 |
| Health (5-25) | 13.1 | (3.2) |  | 12.7 | (3.2) |  | 14.3 | (4.1) |  | 13.5 | (2.9) |  | 13.2 | (2.8) |  | <.001 |
| Role Restriction (7-35) | 19.3 | (5.5) |  | 18.1 | (5.0) |  | 23.5 | (6.6) |  | 20.5 | (5.0) |  | 19.9 | (6.3) |  | <.001 |
| Depression (9-45) | 17.3 | (4.7) |  | 16.7 | (4.8) |  | 19.4 | (5.4) |  | 18.3 | (4.5) |  | 18.3 | (8.8) |  | .19 |
| Spouse/Parenting Partner Relationship (7-35) | 17.9 | (5.9) |  | 16.4 | (4.9) |  | 21.2 | (4.9) |  | 18.2 | (5.1) |  | 17 | (5.2) |  | 0.002 |

NOTE: PSI score ranges are included in parentheses
%: Percentage of respondents who scored above the 85<sup>th</sup> percentile; ImpD: Imprinting Center Defect; UPD: Uniparental Disomy

**Table 4.** Family Quality of Life (FQOL) Scale Scores at Baseline Visit (*n*=214)

| FQOL Score | Class I deletion |  | Class II deletion |  | *UBE3A* pathogenic variant |  | ImpD |  | UPD |  | *p*-value |
| --- | --- | --- | --- | --- | --- | --- | --- | --- | --- | --- | --- |
|  | Mean | (SD) | Mean | (SD) | Mean | (SD) | Mean | (SD) | Mean | (SD) |  |
| **FQOL Total Score** | 4.1 | (0.6) | 4.1 | (0.6) | 3.7 | (0.6) | 3.8 | (0.6) | 4.0 | (0.5) | .10 |
| **Subscale** |  |  |  |  |  |  |  |  |  |  |  |
| Family Interaction | 4.2 | (0.8) | 4.2 | (0.8) | 3.7 | (0.7) | 3.9 | (0.9) | 4.3 | (0.6) | 0.006 |
| Parenting | 4.0 | (0.8) | 4.1 | (0.7) | 3.8 | (0.7) | 4.0 | (0.7) | 4.0 | (0.6) | .54 |
| Emotional Well-Being | 3.5 | (1.0) | 3.5 | (1.1) | 2.8 | (1.1) | 2.8 | (0.9) | 3.1 | (0.7) | 0.005 |
| Physical/Material Well-Being | 4.3 | (0.7) | 4.4 | (0.6) | 4.3 | (0.7) | 4.4 | (0.7) | 4.4 | (0.7) | .71 |
| Disability-Related support | 4.2 | (0.7) | 4.2 | (0.8) | 4.1 | (0.9) | 3.8 | (0.8) | 4.0 | (0.7) | .17 |

NOTE: FQOL scale is scored on a 5-point scale where 1=very dissatisfied, 3=neither satisfied nor dissatisfied, and 5=very satisfied
ImpD: Imprinting Center Defect; UPD: Uniparental Disomy

The impact of molecular subtype on PSI and FQOL scores was explored via GLM and post hoc analysis (Tables 5 and 6, Figures 1-4). Molecular subtype was not a significant predictor of Child Domain stress (Table 5, Figure 1). *UBE3A* pathogenic variant subtype predicted higher Parent Domain (*p_adj_*<.001) and Total Stress (*p_adj_*=.001) compared to the Class II deletion group (Table 5, Figures 2-3). There were no significant differences in FQOL by molecular subtype except for in the Family Interaction subscale where the *UBE3A* pathogenic variant group had lower least-square mean scores than the UPD group (Table 6, Figure 4, *p_adj_*=.033). In within-subtype post hoc comparisons, Emotional Well-Being scores were significantly lower than all other FQOL subscales across subtypes (data not shown).

**Table 5.**
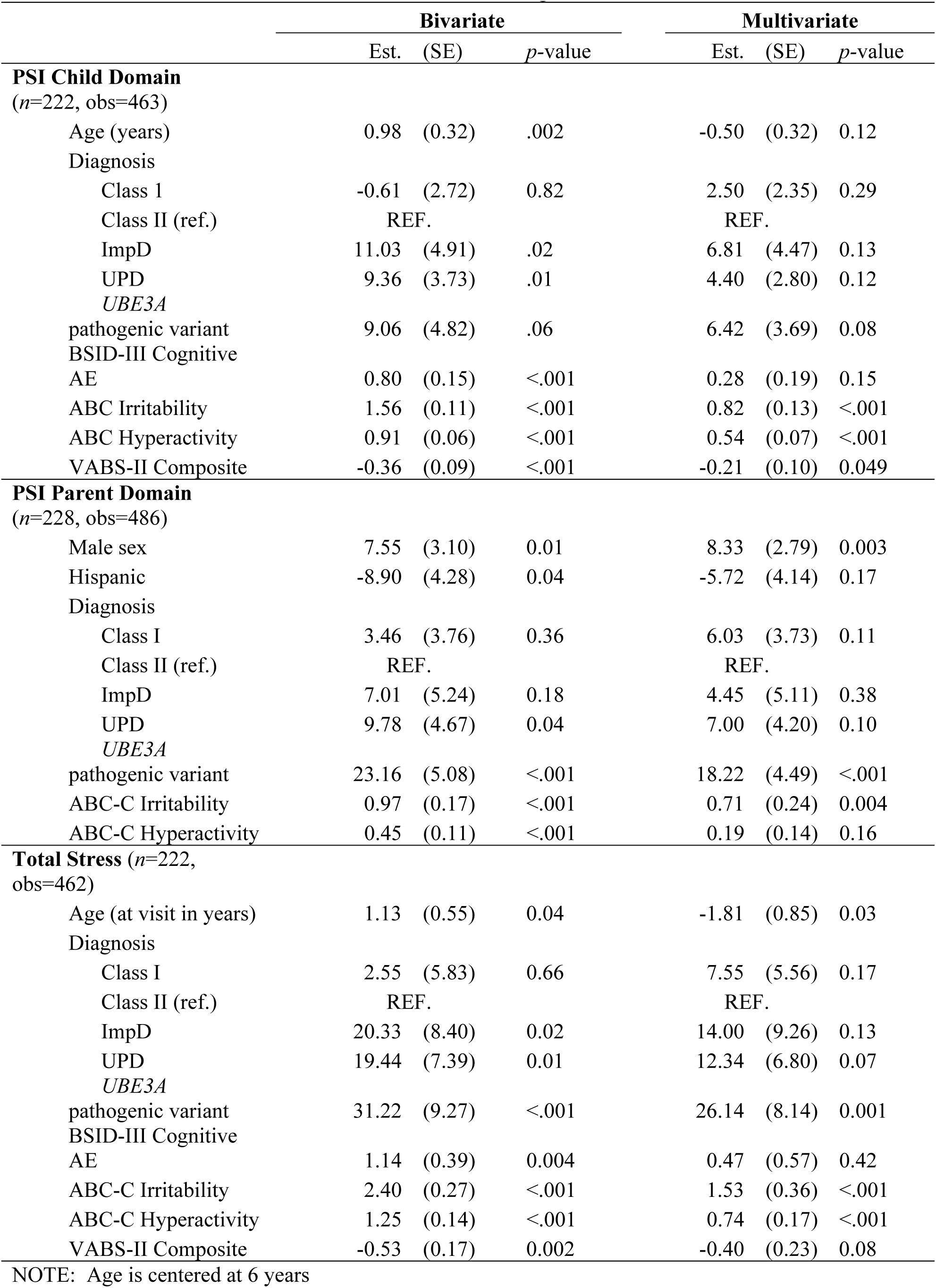
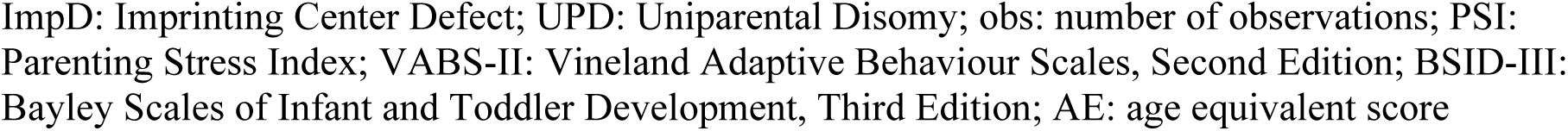
Generalised Linear Models for Predictors of Parenting Stress Index Scores

|  | Bivariate |  |  | Multivariate |  |  |
| --- | --- | --- | --- | --- | --- | --- |
|  | Est. | (SE) | *p*-value | Est. | (SE) | *p*-value |
| **PSI Child Domain** |  |  |  |  |  |  |
| *(n*=222, *obs*=463) |  |  |  |  |  |  |
| Age (years) | 0.98 | (0.32) | .002 | -0.50 | (0.32) | 0.12 |
| Diagnosis |  |  |  |  |  |  |
| Class I | -0.61 | (2.72) | 0.82 | 2.50 | (2.35) | 0.29 |
| Class II (ref.) | REF. |  |  | REF. |  |  |
| ImpD | 11.03 | (4.91) | .02 | 6.81 | (4.47) | 0.13 |
| UPD | 9.36 | (3.73) | .01 | 4.40 | (2.80) | 0.12 |
| *UBE3A* |  |  |  |  |  |  |
| pathogenic variant | 9.06 | (4.82) | .06 | 6.42 | (3.69) | 0.08 |
| BSID-III Cognitive |  |  |  |  |  |  |
| AE | 0.80 | (0.15) | <.001 | 0.28 | (0.19) | 0.15 |
| ABC Irritability | 1.56 | (0.11) | <.001 | 0.82 | (0.13) | <.001 |
| ABC Hyperactivity | 0.91 | (0.06) | <.001 | 0.54 | (0.07) | <.001 |
| VABS-II Composite | -0.36 | (0.09) | <.001 | -0.21 | (0.10) | 0.049 |
| **PSI Parent Domain** |  |  |  |  |  |  |
| *(n*=228, *obs*=486) |  |  |  |  |  |  |
| Male sex | 7.55 | (3.10) | 0.01 | 8.33 | (2.79) | 0.003 |
| Hispanic | -8.90 | (4.28) | 0.04 | -5.72 | (4.14) | 0.17 |
| Diagnosis |  |  |  |  |  |  |
| Class I | 3.46 | (3.76) | 0.36 | 6.03 | (3.73) | 0.11 |
| Class II (ref.) | REF. |  |  | REF. |  |  |
| ImpD | 7.01 | (5.24) | 0.18 | 4.45 | (5.11) | 0.38 |
| UPD | 9.78 | (4.67) | 0.04 | 7.00 | (4.20) | 0.10 |
| *UBE3A* |  |  |  |  |  |  |
| pathogenic variant | 23.16 | (5.08) | <.001 | 18.22 | (4.49) | <.001 |
| ABC-C Irritability | 0.97 | (0.17) | <.001 | 0.71 | (0.24) | 0.004 |
| ABC-C Hyperactivity | 0.45 | (0.11) | <.001 | 0.19 | (0.14) | 0.16 |
| **Total Stress (*n*=222, *obs*=462)** |  |  |  |  |  |  |
| Age (at visit in years) | 1.13 | (0.55) | 0.04 | -1.81 | (0.85) | 0.03 |
| Diagnosis |  |  |  |  |  |  |
| Class I | 2.55 | (5.83) | 0.66 | 7.55 | (5.56) | 0.17 |
| Class II (ref.) | REF. |  |  | REF. |  |  |
| ImpD | 20.33 | (8.40) | 0.02 | 14.00 | (9.26) | 0.13 |
| UPD | 19.44 | (7.39) | 0.01 | 12.34 | (6.80) | 0.07 |
| *UBE3A* |  |  |  |  |  |  |
| pathogenic variant | 31.22 | (9.27) | <.001 | 26.14 | (8.14) | 0.001 |
| BSID-III Cognitive |  |  |  |  |  |  |
| AE | 1.14 | (0.39) | 0.004 | 0.47 | (0.57) | 0.42 |
| ABC-C Irritability | 2.40 | (0.27) | <.001 | 1.53 | (0.36) | <.001 |
| ABC-C Hyperactivity | 1.25 | (0.14) | <.001 | 0.74 | (0.17) | <.001 |
| VABS-II Composite | -0.53 | (0.17) | 0.002 | -0.40 | (0.23) | 0.08 |

NOTE: Age is centered at 6 years
ImpD: Imprinting Center Defect; UPD: Uniparental Disomy; obs: number of observations; PSI: Parenting Stress Index; VABS-II: Vineland Adaptive Behaviour Scales, Second Edition; BSID-III: Bayley Scales of Infant and Toddler Development, Third Edition; AE: age equivalent score

**Table 6.** Generalised Linear Model for Family Quality of Life Scale Predictors with Log Link

|  | Bivariate |  |  | Multivariate |  |  |
| --- | --- | --- | --- | --- | --- | --- |
|  | *Parameter* |  |  | *Parameter* |  |  |
| | Exp($\beta$) | (SE) | *p* | Exp($\beta$) | (SE) | *p* |
| **Model 1: FQOL Family Interaction** (*n*=214, obs=329) |  |  |  |  |  |  |
| Male Sex (ref. is Female) | 0.95 | (0.02) | 0.043 | .97 | (0.018) | 0.11 |
| Diagnosis |  |  |  |  |  |  |
| Class I deletion | 1.02 | (0.029) | 0.60 | 1.02 | (0.023) | .36 |
| Class II deletion (ref.) | REF | REF |  | REF | REF |  |
| *UBE3A* pathogenic variant | 0.86 | (0.038) | <.001 | 0.92 | (0.037) | .04 |
| ImpD | 0.93 | (0.043) | 0.11 | 0.98 | (0.039) | .59 |
| UPD | 1.00 | (0.031) | 0.88 | 1.04 | (0.032) | .17 |
| Child-Domain Stress (in 10 points) | 0.97 | (0.005) | <.001 | 0.99 | (0.007) | 0.13 |
| Parent-Domain Stress (in 10 points) | 0.97 | (0.003) | <.001 | 0.97 | (0.004) | <.001 |
| ABC-C Irritability | 1.00 | (0.002) | 0.02 | 1.00 | (0.002) | 0.84 |
| BSID-III Cognitive AE | 1.00 | (0.001) | 0.01 | 1.00 | (0.001) | 0.38 |
| **Model 2: FQOL Parenting** (*n*=224, obs=351) |  |  |  |  |  |  |
| Male Sex (ref. is Female) | 0.941 | (0.022) | .009 | 0.960 | (0.019) | .04 |
| Child-Domain Stress (in 10 points) | 0.969 | (0.004) | <.001 | 0.985 | (0.005) | .003 |
| Parent-Domain Stress (in 10 points) | 0.967 | (0.003) | <.001 | 0.975 | (0.004) | <.001 |
| **Model 3: FQOL Emotional Well-being** (*n*=209, obs=336) |  |  |  |  |  |  |
| Male Sex (ref. is Female) | 0.896 | (0.037) | 0.008 | 0.900 | (0.032) | .003 |
| Marital Status |  |  |  |  |  |  |
| Single, never married | 1.02 | (0.084) | 0.002 | 1.134 | (0.067) | 0.03 |
| Separated/Divorced | 1.18 | (0.068) | 0.005 | 1.136 | (0.034) | 0.02 |
| Declined | 0.91 | (0.035) | 0.02 | 0.996 | 0.07 | 0.95 |
| Married/Living in marriage-like relationship (ref.) | REF | REF |  | REF | REF |  |
| Diagnosis |  |  |  |  |  |  |
| Class I deletion | 1.026 | (0.050) |  | 1.043 | (0.044) | 0.32 |
| Class II deletion (ref.) | REF | REF |  | REF | REF |  |
| *UBE3A* pathogenic variant | 0.809 | (0.694) | 0.007 | 0.989 | (0.009) | 0.23 |
| ImpD | 0.870 | (0.064) | 0.058 | 0.932 | (0.053) | 0.22 |
| UPD | 0.861 | (0.047) | 0.005 | 0.907 | (0.053) | 0.09 |
| Child-Domain Stress (in 10 points) | 0.967 | (0.008) | <.001 | 0.989 | (0.009) | 0.23 |
| Parent-Domain Stress (in 10 points) | 0.954 | (0.006) | <.001 | 0.966 | (0.007) | <.001 |
| **Model 4: FQOL Physical/Material Well-being** (*n*=201, obs=323) |  |  |  |  |  |  |
| Annual Household Income |  |  |  |  |  |  |
| Under \$25,000 | 0.974 | (0.045) | 0.57 | 0.955 | (0.039) | 0.25 |
| \$25,000-\$49,999 | 0.969 | (0.040) | 0.44 | 0.972 | (0.037) | 0.45 |
| \$50,000-\$99,999 (ref) | REF | REF | | REF | REF | |
| \$100,000 or more | 1.077 | (0.022) | <.001 | 1.060 | (0.019) | .001 |
| Child-Domain Stress (in 10 points) | 0.982 | (0.004) | <.001 | 0.992 | (0.005) | 0.09 |
| Parent-Domain Stress (in 10 points) | 0.980 | (0.003) | <.001 | 0.985 | (0.004) | <.001 |
| **Model 5: FQOL Disability-related Support** (*n*=219, obs=340) |  |  |  |  |  |  |
| Age (in years) | 0.991 | (0.003) | <.001 | 0.992 | (0.004) | .04 |
| Male Sex (ref. is Female) | 0.902 | (0.024) | <.001 | 0.915 | (0.021) | .001 |
| VABS-II Composite (in 5 points) | 1.010 | (0.005) | .04 | 0.992 | (0.006) | 0.21 |
| ABC-C Irritability | 0.993 | (0.002) | <.001 | 1.001 | (0.002) | 0.59 |
| ABC-C Hyperactivity | 0.996 | (0.001) | <.001 | 0.998 | (0.002) | 0.13 |
| Child-Domain Stress (in 10 points) | 0.970 | (0.005) | <.001 | 0.986 | (0.007) | 0.053 |
| Parent-Domain Stress (in 10 points) | 0.978 | (0.004) | <.001 | 0.989 | (0.005) | 0.03 |

NOTE: Effect size in parameter estimate is based on a five-point or ten-point increase in score
NOTE: Exp( $\beta$ )=exponentiated coefficient from a Gamma-generalised linear model (on log scale)
NOTE: Age is centered at 6 years
FQOL: Family Quality of Life; obs: number of observations; ABC-C: Aberrant Behaviour Checklist – Community; ImpD: Imprinting Center Defect; UPD: Uniparental Disomy; VABS-II: Vineland Adaptive Behaviour Scales, Second Edition; BSID-III: Bayley Scales of Infant and Toddler Development, Third Edition; AE: age equivalent score

**Figure 1.**
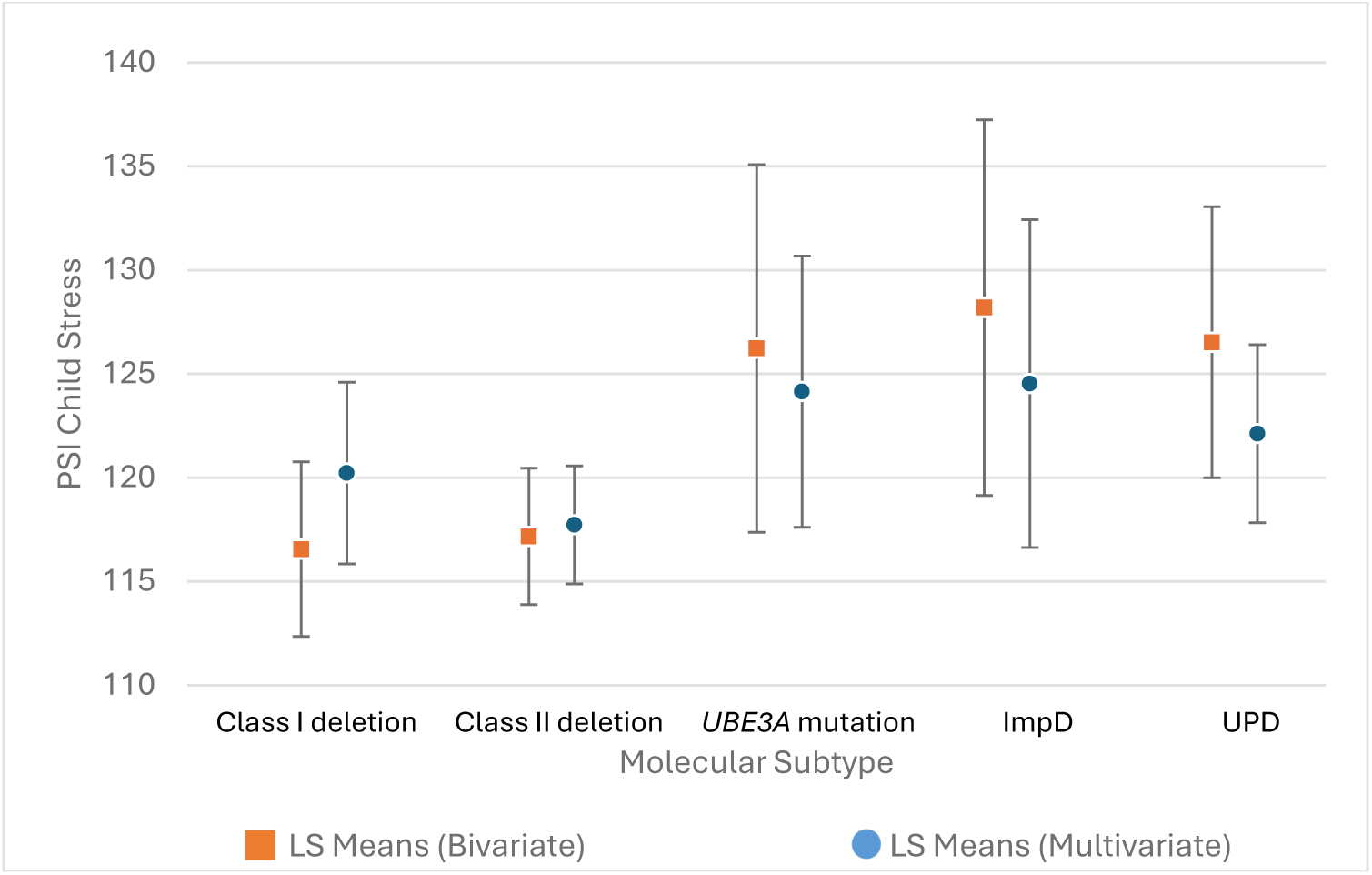
Model-based Parenting Stress Index Child Domain least-square mean scores by molecular subtype. NOTE: Multivariate model controlled for age, BSID-III Cognitive AE, ABC-C Irritability, ABC-C Hyperactivity, and VABS-II Composite with post hoc test of differences corrected for multiple comparisons. Bars indicate significantly different means across groups. PSI: Parenting Stress Index; ImpD: Imprinting Center Defect; UPD: Uniparental Disomy; LS Means: Least-Square Mean Scores; BSID-III: Bayley Scales of Infant and Toddler Development, Third Edition; AE: Age Equivalent; ABC-C: Aberrant Behaviour Checklist – Community; VABS-II: Vineland Adaptive Behaviour Scales, Third Edition

**Figure 2.**
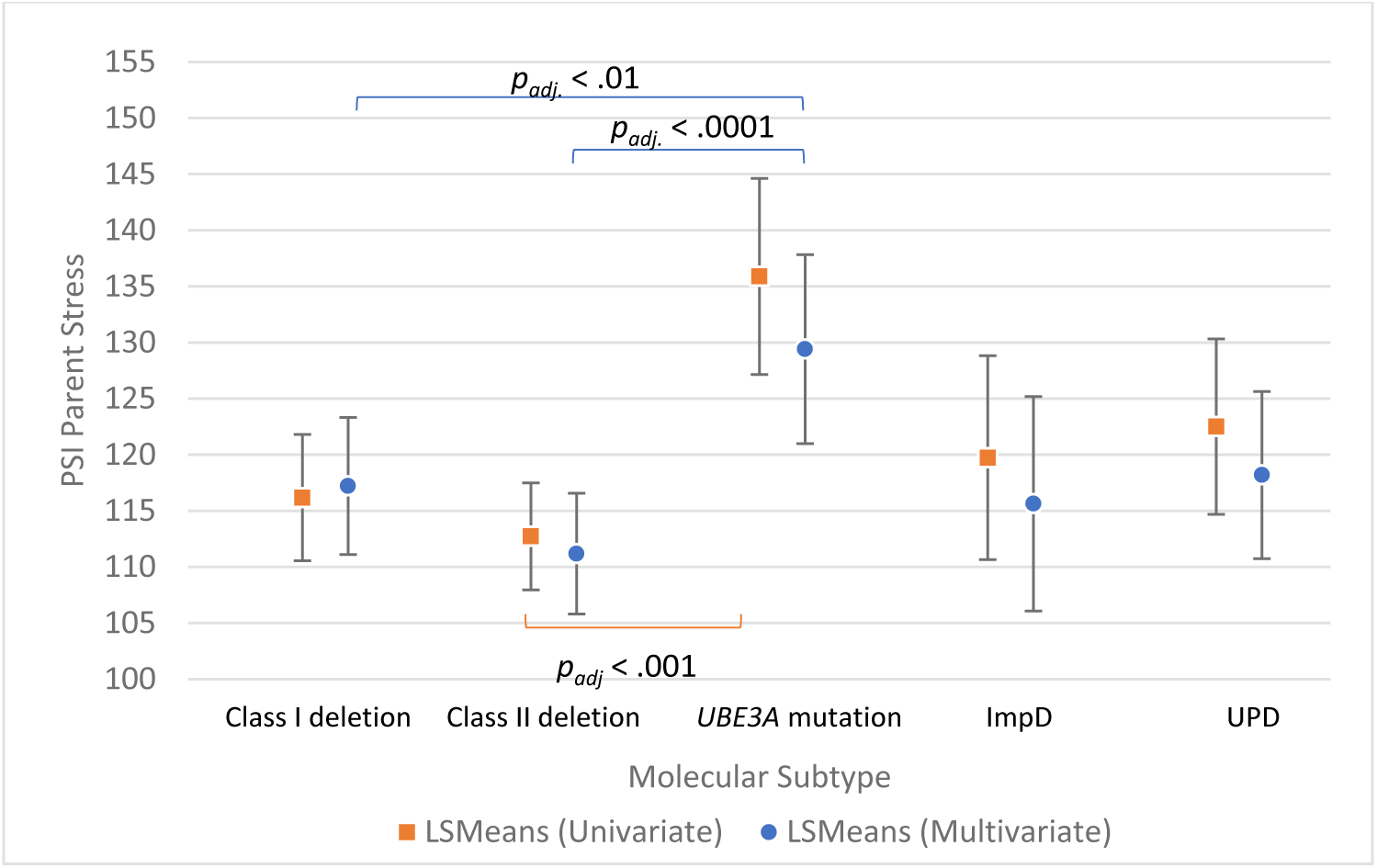
Model-based Parenting Stress Index Parent Domain Least Square-Mean Scores by Molecular Subtype. NOTE: Multivariate model controlling for gender, Hispanic ethnicity, ABC-C Irritability, and ABC-C Hyperactivity with post hoc test of differences corrected for multiple comparisons. Bars indicate significantly different means across groups. PSI: Parenting Stress Index; ImpD: Imprinting Center Defect; UPD: Uniparental Disomy; LS Means: Least-Square Mean Scores

**Figure 3.**
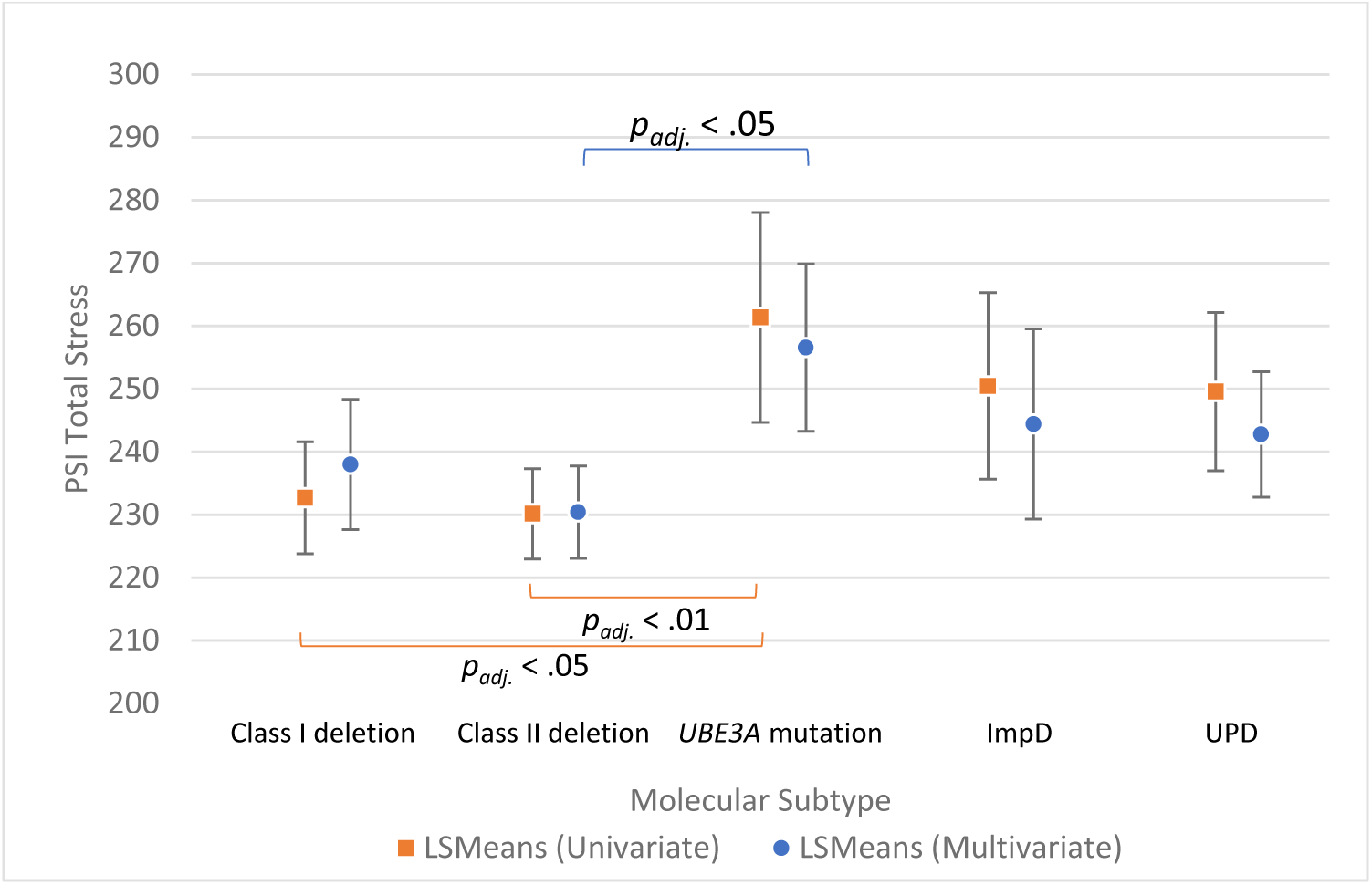
Model-based Parenting Stress Index Total Stress least-square mean scores by molecular subtype. NOTE: Multivariate model controlled for age, gender, BSID-III Cognitive AE, ABC-C Irritability, ABC-C Hyperactivity, and VABS-II Composite with post hoc test of differences correction for multiple comparisons. Bars indicate significantly different means across groups. PSI: Parenting Stress Index; ImpD: Imprinting Center Defect; UPD: Uniparental Disomy; LS Means: Least-Square Mean Scores; BSID-III: Bayley Scales of Infant and Toddler Development, Third Edition; AE: Age Equivalent; ABC-C: Aberrant Behaviour Checklist – Community; VABS-II: Vineland Adaptive Behaviour Scales, Third Edition

**Figure 4.**
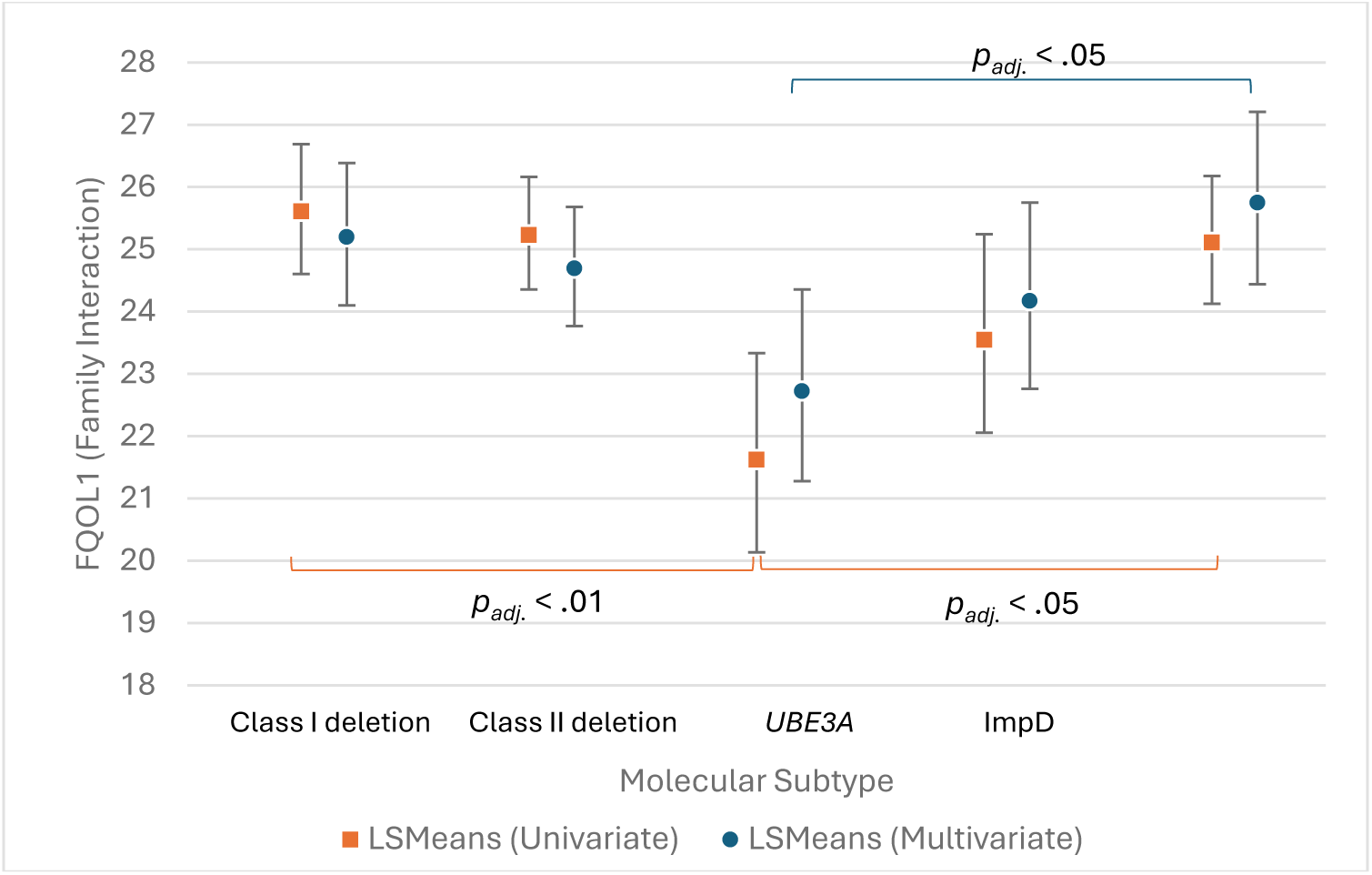
Adjusted least-square mean scores for Family Quality of Life Emotional Well-Being subscale by molecular subtype with 95% confidence limit with post hoc test of differences correction for multiple comparisons. NOTE: Least-square means and confidence intervals for adjusted model controlled for gender, marital status, PSI Child Domain stress, PSI Parent Domain Stress, and were exponentiated for ease of interpretation. Post hoc statistical tests were performed with Bonferroni adjustments. FQOL: Family Quality of Life; ImpD: Imprinting Center Defect; UPD: Uniparental Disomy; LS Means: Least-Square Mean Scores

Changes in parental stress and FQOL over time were explored by analyzing impact of child age on PSI and FQOL scores. Child age was a significant predictor of Total Stress scores. Increasing age was associated with increased Total Stress in the bivariate model (*p*=.04) but was associated with decreased Total Stress in the multivariate model after controlling for sex, subtype, cognitive ability, adaptive functioning, and behaviour (Table 5, *p*=.03). FQOL scores were not significantly affected by child age except in the Disability-Related Support subscale, where satisfaction decreased with increasing child age (Table 6, *p*=.04).

### Predictors of PSI and FQOL scores

ABC-C Irritability scores were significant predictors of Child Domain (p<.001), Parent Domain (*p*=.004), and Total (*p*<.001) stress, while ABC-C Hyperactivity scores also predicted an increase in Child Domain and Total stress (Table 5, *p*<.001). Higher BSID-III Cognitive AE predicted higher levels of Child Domain and Total stress in the bivariate model, but this effect was not significant when controlling for age, sex, behaviour, and adaptive functioning. Higher VABS-II Composite scores predicted lower Child Domain stress in the multivariate model (*p*=.049). Hispanic ethnicity predicted lower Parent Domain stress in the bivariate model, but the interaction was no longer significant in the multivariate model. Other covariates tested were not significant.

Participant sex was a significant predictor of FQOL Parenting, Emotional Well-Being, and Disability-Related Support subscale scores (Table 6). Parents of male individuals scored lower than parents of female individuals in these subscales in the multivariate model. ABC-C Irritability and Hyperactivity scores were not significant predictors of FQOL in the multivariable model. Higher VABS-II Composite scores were associated with increased satisfaction in the Disability-Related Support subscale, but this effect was not significant in the multivariate model. Single and separated/divorced marital status was associated with higher Emotional Well-being subscale scores (*p*=.03 and *p*=.02, respectively). Higher family income was associated with increased Physical/Material Well-Being, with families earning $100,000 or more scoring higher in Physical/Material Well-Being than families earning $50,000-$99,000. Post hoc analysis showed families with annual incomes of $100,000 or more had significantly higher Physical/Material Well-Being scores than those with incomes below $25,000 (*p_adj_* = .041) and $50,000-$99,999 (*p_adj_* = .008) (Figure 5).

**Figure 5.**
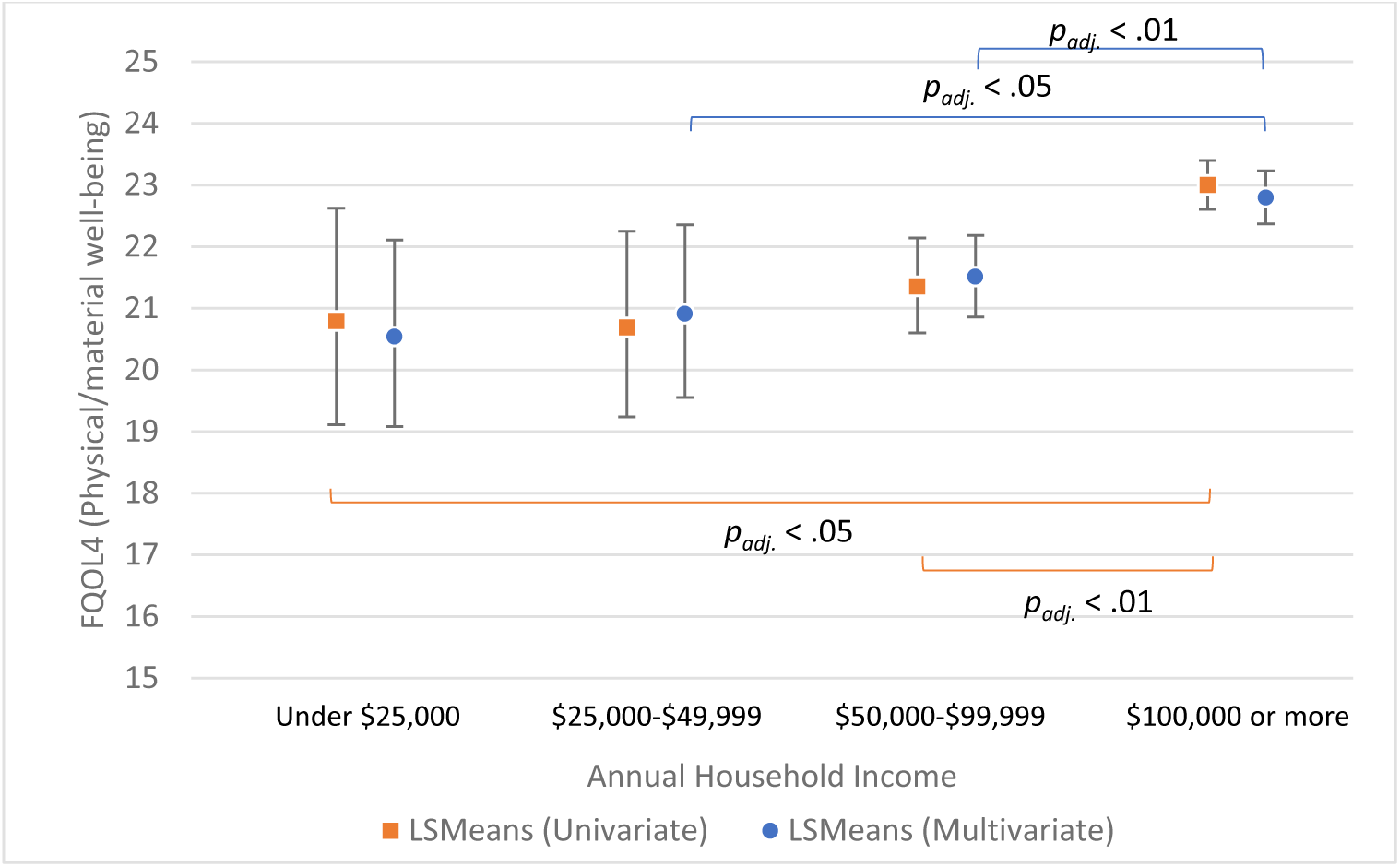
Adjusted mean scores for FQOL Physical/Mental Well-being by Annual Household Income with 95% Confidence Limit with post hoc test of differences correction for multiple comparisons. NOTE: Least-square Means and confidence intervals were adjusted for PSI Child Domain stress and Parent Domain stress as well as for repeated measures and exponentiated for ease of interpretation. Post hoc statistical tests were performed with Bonferroni adjustments. PSI: Parenting Stress Index; FQOL: Family Quality of Life; LS Means: Least-Square Mean Scores

### Impact of parental stress on FQOL

PSI Parent Domain, Child Domain, and Total stress scores were analyzed as predictors of FQOL (Table 6). After controlling for significant covariates, Child Domain stress was associated with a significant decrease in Parenting subscale FQOL satisfaction (*p*=.003). Parent Domain stress was associated with a significant decrease in FQOL satisfaction across all FQOL subscales.

Table 7 outlines associations of the PSI Child and Parent subdomains with FQOL subscales, adjusted for covariates (Table S1). Correlations between PSI and FQOL scores at the baseline visit were first calculated to examine relationships between PSI Domains and FQOL subscales (Table S2). Parent Domain stress was negatively and significantly correlated with FQOL across subtypes, while Child Domain stress was negatively and significantly correlated with FQOL in the deletion groups. Only one significant correlation existed between Child Domain stress and Disability-Related Support satisfaction in the non-deletion groups (Table S2).

**Table 7.**
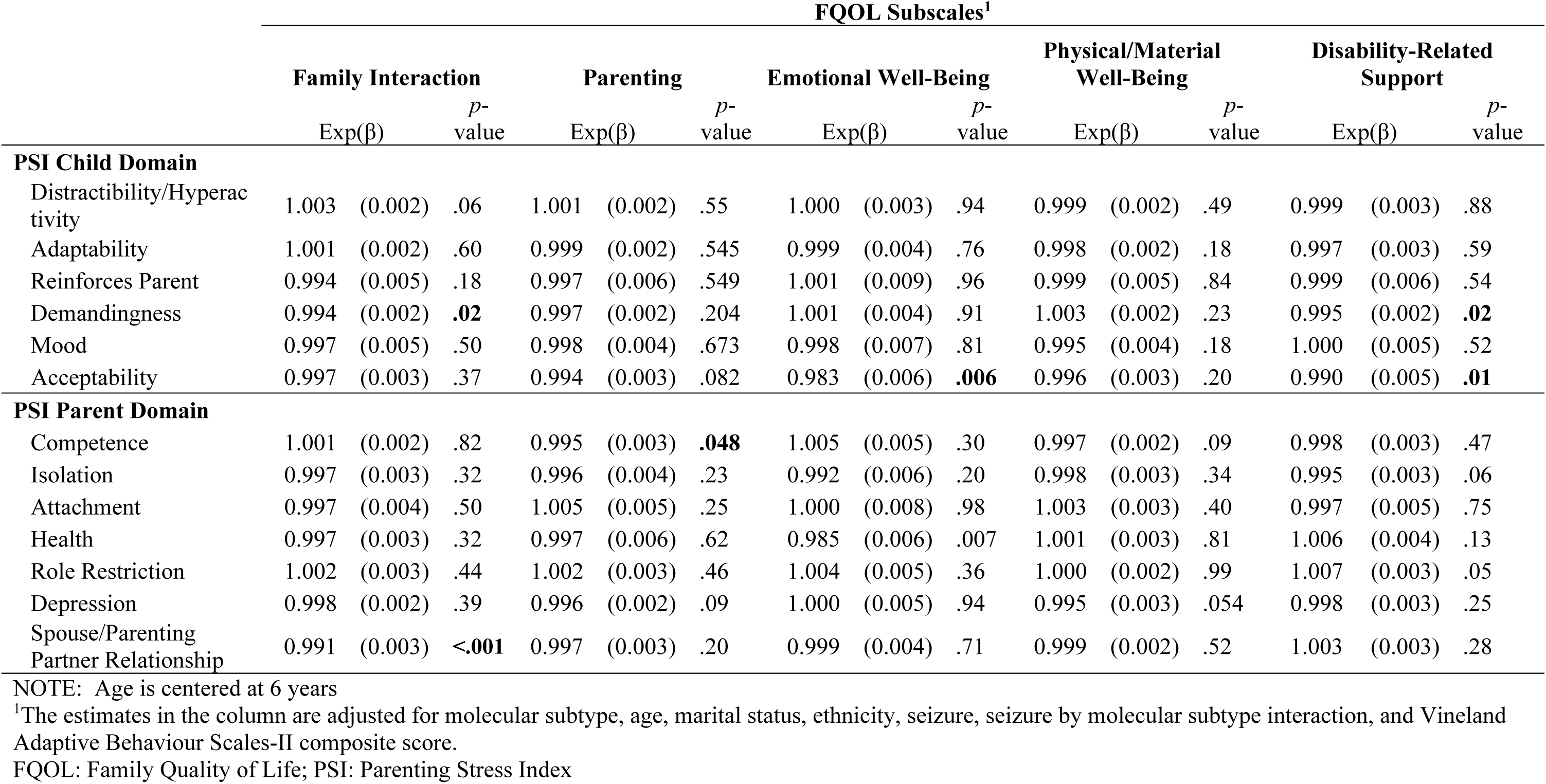
Model-Based Associations Between Parenting Stress Index Domains and Family Quality of Life Subscales (*n*=159)

|  | FQOL Subscales<sup>1</sup> |  |  |  |  |  |  |  |  |  |  |  |  |  |  |
| --- | --- | --- | --- | --- | --- | --- | --- | --- | --- | --- | --- | --- | --- | --- | --- |
|  | Family Interaction |  |  | Parenting |  |  | Emotional Well-Being |  |  | Physical/Material Well-Being |  |  | Disability-Related Support |  |  |
|  | Exp(β) |  | p-value | Exp(β) |  | p-value | Exp(β) |  | p-value | Exp(β) |  | p-value | Exp(β) |  | p-value |
| PSI Child Domain |  |  |  |  |  |  |  |  |  |  |  |  |  |  |  |
| Distractibility/Hyperactivity | 1.003 | (0.002) | .06 | 1.001 | (0.002) | .55 | 1.000 | (0.003) | .94 | 0.999 | (0.002) | .49 | 0.999 | (0.003) | .88 |
| Adaptability | 1.001 | (0.002) | .60 | 0.999 | (0.002) | .545 | 0.999 | (0.004) | .76 | 0.998 | (0.002) | .18 | 0.997 | (0.003) | .59 |
| Reinforces Parent | 0.994 | (0.005) | .18 | 0.997 | (0.006) | .549 | 1.001 | (0.009) | .96 | 0.999 | (0.005) | .84 | 0.999 | (0.006) | .54 |
| Demandingness | 0.994 | (0.002) | **.02** | 0.997 | (0.002) | .204 | 1.001 | (0.004) | .91 | 1.003 | (0.002) | .23 | 0.995 | (0.002) | **.02** |
| Mood | 0.997 | (0.005) | .50 | 0.998 | (0.004) | .673 | 0.998 | (0.007) | .81 | 0.995 | (0.004) | .18 | 1.000 | (0.005) | .52 |
| Acceptability | 0.997 | (0.003) | .37 | 0.994 | (0.003) | .082 | 0.983 | (0.006) | **.006** | 0.996 | (0.003) | .20 | 0.990 | (0.005) | **.01** |
| PSI Parent Domain |  |  |  |  |  |  |  |  |  |  |  |  |  |  |  |
| Competence | 1.001 | (0.002) | .82 | 0.995 | (0.003) | **.048** | 1.005 | (0.005) | .30 | 0.997 | (0.002) | .09 | 0.998 | (0.003) | .47 |
| Isolation | 0.997 | (0.003) | .32 | 0.996 | (0.004) | .23 | 0.992 | (0.006) | .20 | 0.998 | (0.003) | .34 | 0.995 | (0.003) | .06 |
| Attachment | 0.997 | (0.004) | .50 | 1.005 | (0.005) | .25 | 1.000 | (0.008) | .98 | 1.003 | (0.003) | .40 | 0.997 | (0.005) | .75 |
| Health | 0.997 | (0.003) | .32 | 0.997 | (0.006) | .62 | 0.985 | (0.006) | .007 | 1.001 | (0.003) | .81 | 1.006 | (0.004) | .13 |
| Role Restriction | 1.002 | (0.003) | .44 | 1.002 | (0.003) | .46 | 1.004 | (0.005) | .36 | 1.000 | (0.002) | .99 | 1.007 | (0.003) | .05 |
| Depression | 0.998 | (0.002) | .39 | 0.996 | (0.002) | .09 | 1.000 | (0.005) | .94 | 0.995 | (0.003) | .054 | 0.998 | (0.003) | .25 |
| Spouse/Parenting Partner Relationship | 0.991 | (0.003) | **<.001** | 0.997 | (0.003) | .20 | 0.999 | (0.004) | .71 | 0.999 | (0.002) | .52 | 1.003 | (0.003) | .28 |

NOTE: Age is centered at 6 years
<sup>1</sup>The estimates in the column are adjusted for molecular subtype, age, marital status, ethnicity, seizure, seizure by molecular subtype interaction, and Vineland Adaptive Behaviour Scales-II composite score.
FQOL: Family Quality of Life; PSI: Parenting Stress Index

After controlling for covariates, only a few associations between PSI subdomain scores and FQOL subscale scores remained statistically significant (Table 7). Higher Child Demandingness (PSI Child Domain) and Spouse/Parenting Partner Relationship (PSI Parent Domain) scores were associated with decreased Family Interaction FQOL scores; higher Competence (PSI Parent Domain) scores, indicating lower feelings of parental competence, had negative association with Parenting FQOL scores; increased Acceptability (PSI Child Domain) scores, indicating differences between parent expectation of child abilities and their perceived actual abilities, and increased Health (PSI Parent Domain) scores, indicating stress related to parent health, had an inverse relationship with Emotional Well-Being FQOL scores; and increased Demandingness and Acceptability (PSI Child Domain) scores had a negative association with Disability-Related Support FQOL scores (Table 7).

## DISCUSSION

We aimed to describe parental stress and FQOL across AS molecular subtypes and the factors that contributed to each. Parental stress was lower in parents of individuals with deletion subtypes compared to those with non-deletion subtypes, as previously demonstrated using a partially overlapping dataset.^13^ This is the first study to describe parenting stress in parents of individuals with *UBE3A* pathogenic variants; we found that this group experienced the highest rates of clinically significant stress, driven disproportionately by parent factors compared to other subtypes.^21^ Nineteen to 48% of participants reported clinically significant parental stress depending on molecular subtype group, indicating need for interventions to reduce stress.

FQOL satisfaction reported here is similar to previously reported levels in families with children with ID.^22–24^ Families experience lowest satisfaction with their support systems and ability to pursue independent interests. Parents of children with ID often face multiple demands in their caregiving role (managing financial stress, coordinating care, emotional stress, and social isolation).^35^ These factors likely contribute to lower satisfaction in support systems and time for self in AS families. Resources that foster social support and activities outside the parenting role may improve emotional well-being.^36^

As participants aged, we saw a small decrease in stress but also in Disability-Related Support satisfaction. Older age is associated with lower health-related QoL, possibly due to increased health risks, anxiety, and divergence from developmental level of peers amplifying perception of deficits.^17,18^ These effects may explain decreased FQOL associated with increased age. Reduction in parental stress over time may be due to families developing support systems and coping strategies which improve parent well-being, marital adjustment, and positive child-parent relationships over time.^37^

Maladaptive behaviours correlated with increased parental stress but did not strongly impact FQOL, consistent with previous research.^5,17,38^ Irritability may strain the parent-child relationship and increase parental stress.^21^ Irritability additionally contributed to parent-related stressors such as perceptions of competence, attachment, well-being, and relationships. The relationship between parental stress and maladaptive behaviours may be bidirectional, with increased stress leading to erosion of the parent-child relationship and contributing to behavioural problems.^39,40^ Additional studies may seek to examine how behaviour affects stress by analyzing the effects of behaviour on specific PSI subdomains.

Families with male children had higher stress and lower FQOL than families with female children, possibly due to increased rates of aggressive behaviours (e.g., hair-pulling, biting) which occur more frequently in male individuals with AS.^5,41^ Families with higher incomes were more satisfied with their material and physical well-being, reflecting the importance of income to meet basic needs such as transportation, healthcare, and feeling safe.^22^

Medical complexity of the individual with AS, indicated by seizure severity and hospitalizations per year, as well as sleep difficulties, did not affect stress or FQOL. Individuals with AS have high health burden, which harms individual QoL.^42,43,18,19^ Epilepsy severity and hospitalization can cause increased parental stress,^44,45^ but our results suggest these are not sources of long-term stress in AS. Sleep difficulties in the child did not affect stress, inconsistent with previous research.^14,17^ However, we did not utilise a standardised sleep questionnaire or objective sleep assessment, warranting future research on this association. Cognitive functioning did not affect parental stress or FQOL. Improved adaptive functioning had a minimal positive effect on Child Domain stress.

Hagenaar et al. (2024) reported child characteristics associated with health-related QoL and parental stress in a sample of 73 children with AS and did not find significant differences in stress based on molecular subtype, epilepsy, sleep problems, or cognitive function (measured by BSID-III), but did report high impact of child sleep problems on parent emotional-and time-burden.^17^ Our study supports these results by further describing stress across molecular subtype in a larger sample, as well as describing the effects of age on stress assessed across multiple visits. Finally, we contribute to QoL literature by describing FQOL across AS subtypes.

Parental stress and family quality of life are closely related, and this was reflected in our results.^25,28^ Higher levels of parental stress predicted a small decrease in FQOL. These effects were not previously studied in AS, but prior ID research has shown similar associations.^25,46^ Stress-FQOL associations were driven by parent-related factors, such as health concerns and feelings of incompetence, as well as child-related factors, such as child demandingness. Lower satisfaction with outside supports and with the parenting role can be caused by and contribute to lower feelings of parental competence.^47^ Parents who experience high demands for attention, high defiance and aggression, and discrepancies in expectations versus realities of child behaviours and abilities may be less satisfied with their family cohesion.^21^

Interventions that reduce parental stress may directly improve parent well-being and support family interactions, as well as indirectly support effective parenting strategies and foster positive outcomes for children with IDs.^36,39,48^ Research suggests that referral to professional counseling would be recommended for the 19-48% of families in our sample who reached the clinical stress cut-off,^21^ but actual rates of counseling in our study population are not known.

Other interventions such as respite care, case management involvement, group cognitive behavioural therapy, and parent-led support groups may be effective in lowering parental stress in AS families.^36,48^ Treatments that target challenging child behaviours may also reduce parental stress.^5,14^ We identified molecular subtype, child sex, and family income as predictors of parental stress and FQOL. While these are not clinically modifiable, clinicians should be mindful of these factors to identify families who may be at increased risk for stress and family dysfunction.

Clinicians who serve individuals with AS and their families are in an ideal position to understand the demands placed on parents, identify families at increased risk for elevated stress, and help initiate interventions to reduce parental stress and maximise family well-being.

### Limitations

There are several limitations to this study. Firstly, the sample lacked racial and socioeconomic diversity. Only 7% of participants reported income below $25,000, and only 9% reported non-White race. Families of children with and without IDs from groups who are marginalised, including non-White, Hispanic, and low-income families, experience disparities in health outcomes and access, and may be variably vulnerable to parental stress and its effects on FQOL, depending on structural and cultural factors.^16,49,50^ Additionally, participants in this study likely had ready access to resources which allowed them to participate in a time-intensive study which may not reflect access in the general population. Barriers exist to marginalised groups participating in research.^51^ There are no studies that focus on parental stress and FQOL in underrepresented families of individuals living with AS. Future AS studies should expand their reach to explore how parental stress and FQOL manifest in non-White and lower-income families to support social and cultural competence in assessing and responding to these outcomes.

Secondly, assessment of parental stress and FQOL had limitations. Elevated stress may not indicate elevated *distress*. Child Domain stress was less strongly associated with FQOL than Parent Domain stress despite higher Child Domain compared to Parent Domain stress levels.

This suggests that child characteristics evaluated on the PSI may not represent stressful characteristics in these families due to resilience and positive coping.^37,52^ The PSI does not capture the positive aspects of the parent-child relationship; however, the FQOL scale aims to capture positive aspects of raising a child with ID.^32^ Having a child with AS has diverse impacts on families, and identifying positive aspects of parent-child relationships may elucidate how families adapt to challenges. The Beach FQOL Scale was designed to assess outcomes of services for people with IDs and may not encompass all aspects of FQOL that families find important.

Only one primary caregiver completed the PSI and FQOL, limiting the perspectives of parental stress and FQOL to one parenting partner. Mothers and fathers respond to parenting a child with health problems differently,^53,54^ but we lacked sufficient data to analyze stress in mothers and fathers independently. Future studies should collect PSI data from all parent partners, and FQOL data from multiple members of the immediate family, to learn how different family members perceive stress and family well-being.

Finally, we collected data at two to three time points for each participant on average, limiting our assessment of how parental stress or FQOL change over the course of childhood. Longitudinal studies of parental stress and FQOL over many years may be pursued to further this understanding.

## Conclusions

Parental stress is elevated but decreases over time and is highest in families of individuals living with *UBE3A* pathogenic variants. Maladaptive behaviours and child male sex are associated with higher parental stress. FQOL satisfaction was generally similar across molecular subtypes and was stable over time. Stress had a small negative impact on FQOL. Interventions that reduce parental stress may improve parent well-being and support family interactions.

## Acknowledgements

This study was supported by NIH U54 RR019478 (awarded to Arthur L. Beaudet) from the National Center for Research Resources (NCRR) and NIH U54 HD061222 (awarded to Alan Percy) from the National Institute of Child Health and Human Development (NICHD), both components of the National Institutes of Health (NIH). We would like to thank our study participants and their families for their continued commitment to this longitudinal study. We would also like to thank all the site principal investigators and coordinators who facilitated the collection of these data.

## Data availability statement

The data that support the findings of this study are available from the corresponding author upon reasonable request.

## REFERENCES

1. Bird LM. Angelman syndrome: review of clinical and molecular aspects. Application of clinical genetics. 2014;7:93–104.

2. Williams CA, Beaudet AL, Clayton-Smith J, et al. Angelman syndrome 2005: Updated consensus for diagnostic criteria. American journal of medical genetics Part A. 2006;140A(5):413–418.

3. Sadhwani A, Wheeler A, Gwaltney A, et al. Developmental Skills of Individuals with Angelman Syndrome Assessed Using the Bayley-III. J Autism Dev Disord. 2023;53(2):720–737.

4. Clarke DJ, Marston G. Problem behaviors associated with 15q-Angelman syndrome. American journal of mental retardation. 2000;105(1):25–31.

5. Sadhwani A, Willen JM, LaVallee N, et al. Maladaptive behaviors in individuals with Angelman syndrome. American journal of medical genetics Part A. 2019;179(6):983–992.

6. Pelc K, Cheron G, Boyd SG, Dan B. Are there distinctive sleep problems in Angelman syndrome? Sleep medicine. 2007;9(4):434–441.

7. Wheeler AC, Sacco P, Cabo R. Unmet clinical needs and burden in Angelman syndrome: a review of the literature. Orphanet journal of rare diseases. 2017;12(1):164–164.

8. Keary CJ, McDougle CJ. Current and emerging treatment options for Angelman syndrome. Expert Rev Neurother. 2023;23(9):835–844.

9. Griffith GM, Hastings RP, Oliver C, et al. Psychological well-being in parents of children with Angelman, Cornelia de Lange and Cri du Chat syndromes. Journal of intellectual disability research. 2011;55(4):397–410.

10. Thomson A, Glasson E, Roberts P, Bittles A. “Over time it just becomes easier…”: parents of people with Angelman syndrome and Prader-Willi syndrome speak about their carer role. Disability and rehabilitation. 2017;39(8):763–770.

11. van den Borne HW, van Hooren RH, van Gestel M, Rienmeijer P, Fryns JP, Curfs LMG. Psychosocial problems, coping strategies, and the need for information of parents of children with Prader–Willi syndrome and Angelman syndrome. Patient education and counseling. 1999;38(3):205–216.

12. Wulffaert J, Scholte EM, Van Berckelaer-Onnes IA. Maternal parenting stress in families with a child with Angelman syndrome or Prader-Willi syndrome. Journal of intellectual & developmental disability. 2010;35(3):165–174.

13. Miodrag N, Peters S. Parent stress across molecular subtypes of children with Angelman syndrome: Parent stress across molecular subtypes. Journal of intellectual disability research. 2015;59(9):816–826.

14. Goldman SE, Bichell TJ, Surdyka K, Malow BA. Sleep in children and adolescents with Angelman syndrome: association with parent sleep and stress. Journal of intellectual disability research. 2012;56(6):600–608.

15. Fang Y, Luo J, Boele M, Windhorst D, van Grieken A, Raat H. Parent, child, and situational factors associated with parenting stress: a systematic review. Eur Child Adolesc Psychiatry. 2024;33(6):1687–1705.

16. Cardoso JB, Padilla YC, Sampson M. Racial and ethnic variation in the predictors of maternal parenting stress. J Soc Serv Res. 2010;36(5):429–444.

17. Hagenaar DA, Bindels-de Heus K, Lubbers K, et al. Child characteristics associated with child quality of life and parenting stress in Angelman syndrome. J Intellect Disabil Res. 2024;68(3):248–263.

18. Xia NY, Grant ML, Benjamin NL, Valencia I. Quality of Life in Angelman Syndrome: A Caregivers’ Survey. Pediatr Neurol. 2023;149:19–25.

19. Khan N, Cabo R, Burdine RD, et al. Health-related quality of life and medication use among individuals with Angelman syndrome. Qual Life Res. 2023;32(7):2059–2067.

20. 603 CMR 28:00 Special Education. In: Education DoEaS, ed2022.

21. Abidin RR. Parenting Stress Index: Manual, 3rd edn. Psychological Assessment Resources, Inc, Odessa, FL. 1995.

22. Lesa H, Janet M, Denise P, Jean Ann S, Ann T. Assessing Family Outcomes: Psychometric Evaluation of the Beach Center Family Quality of Life Scale. Journal of marriage and family. 2006;68(4):1069–1083.

23. Boehm TL, Carter EW, Taylor JL. Family Quality of Life During the Transition to Adulthood for Individuals With Intellectual Disability and/or Autism Spectrum Disorders. American Journal on Intellectual and Developmental Disabilities. 2015;120(5):395–411.

24. Staunton E, Kehoe C, Sharkey L. Families under pressure: stress and quality of life in parents of children with an intellectual disability. Irish Journal of Psychological Medicine. 2020:1–8.

25. Jenaro C, Flores N, Gutiérrez-Bermejo B, Vega V, Pérez C, Cruz M. Parental Stress and Family Quality of Life: Surveying Family Members of Persons with Intellectual Disabilities. Int J Environ Res Public Health. 2020;17(23).

26. Werner S, Edwards M, Baum N, Brown I, Brown RI, Isaacs BJ. Family quality of life among families with a member who has an intellectual disability: an exploratory examination of key domains and dimensions of the revised FQOL Survey. J Intellect Disabil Res. 2009;53(6):501–511.

27. Fong VC, Gardiner E, Iarocci G. Can a combination of mental health services and ADL therapies improve quality of life in families of children with autism spectrum disorder? Quality of life research. 2020;29(8):2161–2170.

28. Hsiao Y-J, Higgins K, Pierce T, Whitby PJS, Tandy RD. Parental stress, family quality of life, and family-teacher partnerships: Families of children with autism spectrum disorder. Research in developmental disabilities. 2017;70:152–162.

29. Bayley N. Bayley Scales of Infant and Toddler Development, Third Edition (Bayley-III). *Harcourt Assessment*, Inc. 2005.

30. Sparrow SS, Cicchettim DV, Balla DA. Vineland Adaptive Behavior Scales, Second Edition (Vineland-II). APA PsychTests. 2005.

31. Aman MG, Singh NN, Stewart AW, Field CJ. The aberrant behavior checklist: a behavior rating scale for the assessment of treatment effects. American journal of mental deficiency. 1985;89(5):485.

32. Summers JA, Poston DJ, Turnbull AP, et al. Conceptualizing and measuring family quality of life. J Intellect Disabil Res. 2005;49(Pt 10):777–783.

33. Thibert RL, Conant KD, Braun EK, et al. Epilepsy in Angelman syndrome: a questionnaire-based assessment of the natural history and current treatment options. Epilepsia. 2009;50(11):2369–2376.

34. Gentile JK, Tan W-H, Horowitz LT, et al. A Neurodevelopmental Survey of Angelman Syndrome With Genotype-Phenotype Correlations. Journal of developmental and behavioral pediatrics. 2010;31(7):592–601.

35. Thompson R, Kerr M, Glynn M, Linehan C. Caring for a family member with intellectual disability and epilepsy: Practical, social and emotional perspectives. Seizure. 2014;23(10):856–863.

36. Miranda A, Mira A, Berenguer C, Rosello B, Baixauli I. Parenting Stress in Mothers of Children With Autism Without Intellectual Disability. Mediation of Behavioral Problems and Coping Strategies. Front Psychol. 2019;10:464.

37. Gerstein ED, Crnic KA, Blacher J, Baker BL. Resilience and the course of daily parenting stress in families of young children with intellectual disabilities. Journal of Intellectual Disability Research. 2009;53(12):981–997.

38. Tomanik S, Harris GE, Hawkins J. The relationship between behaviours exhibited by children with autism and maternal stress. Journal of Intellectual & Developmental Disability. 2004;29(1):16–26.

39. Baker BL, McIntyre LL, Blacher J, Crnic K, Edelbrock C, Low C. Pre-school children with and without developmental delay: behaviour problems and parenting stress over time. J Intellect Disabil Res. 2003;47(Pt 4-5):217–230.

40. Neece CL, Green SA, Baker BL. Parenting stress and child behavior problems: a transactional relationship across time. Am J Intellect Dev Disabil. 2012;117(1):48–66.

41. Barroso NE, Mendez L, Graziano PA, Bagner DM. Parenting Stress through the Lens of Different Clinical Groups: a Systematic Review & Meta-Analysis. Journal of Abnormal Child Psychology. 2018;46(3):449–461.

42. Khan N, Cabo R, Tan WH, Tayag R, Bird LM. Healthcare burden among individuals with Angelman syndrome: Findings from the Angelman Syndrome Natural History Study. Molecular genetics & genomic medicine. 2019;7(7):e00734-n/a.

43. Khan N, Cabo R, Tan W-H, Tayag R, Bird LM. An observational study of pediatric healthcare burden in Angelman syndrome: results from a real-world study. Orphanet journal of rare diseases. 2019;14(1):239–239.

44. Zdun-Ryżewska A, Nadrowska N, Błażek M, Białek K, Zach E, Krywda-Rybska D. Parent’s Stress Predictors during a Child’s Hospitalization. Int J Environ Res Public Health. 2021;18(22).

45. Operto FF, Mazza R, Pastorino GMG, Campanozzi S, Verrotti A, Coppola G. Parental stress in a sample of children with epilepsy. Acta Neurol Scand. 2019;140(2):87–92.

46. Staunton E, Kehoe C, Sharkey L. Families under pressure: stress and quality of life in parents of children with an intellectual disability. Ir J Psychol Med. 2020:1–8.

47. Jandrić S, Kurtović A. Parenting Sense of Competence in Parents of Children With and Without Intellectual Disability. Eur J Psychol. 2021;17(2):75–91.

48. Hastings RP, Beck A. Practitioner review: stress intervention for parents of children with intellectual disabilities. J Child Psychol Psychiatry. 2004;45(8):1338–1349.

49. Nomaguchi K, House AN. Racial-ethnic disparities in maternal parenting stress: the role of structural disadvantages and parenting values. J Health Soc Behav. 2013;54(3):386–404.

50. Iadarola S, Pérez-Ramos J, Smith T, Dozier A. Understanding stress in parents of children with autism spectrum disorder: A focus on under-represented families. Int J Dev Disabil. 2019;65(1):20–30.

51. Heller C, Balls-Berry JE, Nery JD, et al. Strategies addressing barriers to clinical trial enrollment of underrepresented populations: A systematic review. Contemporary Clinical Trials. 2014;39(2):169–182.

52. Peer JW, Hillman SB. Stress and Resilience for Parents of Children With Intellectual and Developmental Disabilities: A Review of Key Factors and Recommendations for Practitioners. Journal of Policy and Practice in Intellectual Disabilities. 2014;11(2):92–98.

53. Pelchat D, Lefebvre H, Levert M-J. Gender differences and simililarities in the experience of parenting a child with a health problem: current state of knowledge. Journal of Child Health Care. 2007;11(2):112–131.

54. Potter SN, Harvey DJ, Sterling A, Abbeduto L. Mental Health Challenges, Parenting Stress, and Features of the Couple Relationship in Parents of Children With Fragile X Syndrome. Front Psychiatry. 2022;13:857633.

