## Supplemental Tables S1 and S2 for "Parental stress and family quality of life in families of individuals living with Angelman syndrome"

Table S1. Test of Fixed Effects of Model Controls (*n*=159)

|  | **FQOL Subscales** | | | | |
| --- | --- | --- | --- | --- | --- |
|  | Family Interaction | Parenting | Emotional Well-Being | Physical/Material Well-Being | Disability-Related Support |
| Obs | obs=243 | Obs=241 | obs=243 | obs=243 | obs=243 |
| Molecular Subtype (ref=Class II) |  |  |  |  |  |
| Class I deletion | NS | NS | NS | NS | NS |
| ImpD | NS | NS | NS | NS | NS |
| *UBE3A* mutation | NS | NS | NS | NS | NS |
| UPD | NS | NS | NS | NS | NS |
| Age | NS | NS | NS | NS | NS |
| Ethnicity (ref. non-Hispanic) | NS | 0.915 (0.032), *p* = .01 | NS | 0.928 (0.035), *p* = .046 | NS |
| Marital Status | NS | NS | NS | NS | NS |
| Seizure | NS | NS | NS | NS | NS |
| VABS-II Composite | NS | NS | NS | NS | NS |
| Molecular subtype x Seizure | NS | NS | NS | NS | NS |
| NOTE: Age is centered at 6 years and significant results are in exponentiated log  Exp(β) and *p*-value for significant covariates shown.  FQOL: Family Quality of Life; ImpD: Imprinting Center Defect; UPD: Uniparental Disomy; VABS-II: Vineland Adaptive Behaviour Scales, Second Edition; NS: Non-significant | | | | | |

Table S2. Correlations of Family Quality of Life Subscales with Child Domain, Parent Domain, and Total Parenting Stress Index Scores by Molecular Subtype at Baseline Visit (*n*=231)

|  | **PSI Child Domain** | | | | | | | | | |
| --- | --- | --- | --- | --- | --- | --- | --- | --- | --- | --- |
| **FQOL** | Class I deletion | | Class II deletion | | *UBE3A* mutation | | ImpD | | UPD | |
|  | ρ | *p*-value | ρ | *p*-value | ρ | *p*-value | ρ | *p*-value | ρ | *p*-value |
| Family Interaction | -0.39 | .004 | -0.30 | .007 | -0.29 | .16 | -0.29 | .24 | -0.28 | .17 |
| Parenting | -0.34 | .01 | -0.39 | <.001 | -0.40 | .05 | -0.18 | .45 | -0.35 | .08 |
| Emotional Well-Being | -0.17 | .23 | -0.34 | .003 | -0.34 | .10 | -0.26 | .29 | 0.01 | .95 |
| Physical/Material Well-Being | -0.35 | .009 | -0.18 | .12 | -0.34 | .10 | -0.14 | .57 | -0.36 | .08 |
| Disability-Related Support | -0.23 | .09 | -0.33 | .003 | -0.13 | .55 | 0.06 | .81 | -0.54 | .006 |

|  | **PSI Parent Domain** | | | | | | | | | |
| --- | --- | --- | --- | --- | --- | --- | --- | --- | --- | --- |
| **FQOL** | Class I deletion | | Class II deletion | | *UBE3A* mutation | | ImpD | | UPD | |
|  | ρ | *p*-value | ρ | *p*-value | ρ | *p*-value | ρ | *p*-value | ρ | *p*-value |
| Family Interaction | -0.55 | <.001 | -0.46 | <.001 | -0.58 | .003 | -0.63 | .004 | -0.17 | .40 |
| Parenting | -0.36 | .01 | -0.52 | <.001 | -0.46 | .02 | -0.61 | .005 | -0.36 | .07 |
| Emotional Well-Being | -0.35 | .01 | -0.48 | <.001 | -0.52 | .01 | -0.82 | <.001 | -0.18 | .39 |
| Physical/Material Well-Being | -0.33 | .01 | -0.34 | .003 | -0.53 | .007 | -0.42 | .07 | -0.67 | <.001 |
| Disability-Related Support | -0.40 | .003 | -0.31 | .007 | -0.16 | .45 | -0.03 | .90 | -0.64 | <.001 |

|  | | | **PSI Total Stress Score** | | | | | | | | | |
| --- | --- | --- | --- | --- | --- | --- | --- | --- | --- | --- | --- | --- |
| **FQOL** | | | Class I deletion | | Class II deletion | | *UBE3A* mutation | | ImpD | | UPD | |
|  | | | ρ | *p*-value | ρ | *p*-value | ρ | *p*-value | ρ | *p*-value | ρ | *p*-value |
| Family Interaction | | | -0.55 | <.001 | -0.44 | <.001 | -0.48 | .02 | -0.57 | .01 | -0.25 | .23 |
| Parenting | | | -0.41 | .002 | -0.51 | <.001 | -0.47 | .02 | -0.51 | .03 | -0.41 | .04 |
| Emotional Well-Being | | | -0.31 | .02 | -0.48 | <.001 | -0.47 | .02 | -0.69 | .001 | -0.08 | .69 |
| Physical/Material Well-Being | | | -0.39 | .003 | -0.31 | .007 | -0.48 | .02 | -0.36 | .13 | -0.64 | <.001 |
| Disability-Related Support | | | -0.38 | .005 | -0.36 | .001 | -0.16 | .46 | 0.01 | .97 | -0.72 | <.001 |
| Pearson Correlation | | | | | | | | | | | | |
|  |  | -0.1 | | | | | | | | | | |
|  |  | -0.2 | | | | | | | | | | |
|  |  | -0.3 | | | | | | | | | | |
|  |  | -0.4 | | | | | | | | | | |
|  |  | -0.5 | | | | | | | | | | |
|  |  | -0.6 | | | | | | | | | | |
|  |  | -0.7 | | | | | | | | | | |
|  |  | -0.8 | | | | | | | | | | |
| ImpD: Imprinting Center Defect; UPD: Uniparental Disomy | | | | | | | | | | | | |
